# Cerebellum-ventral tegmental connectivity as a mechanism-informed target for apathy in schizophrenia

**DOI:** 10.64898/2026.05.20.26353653

**Authors:** Thomas A.W. Bolton, Lorina Sinanaj, Halil A. Velioglu, Dimitri Van De Ville, Stefan Kaiser, Hengyi Cao, Indrit Bègue

## Abstract

Apathy is a leading driver of functional disability in schizophrenia, yet effective mechanism-based therapies are lacking. We evaluated whether cerebellum–ventral tegmental area functional connectivity (CB-VTA FC) meets the criteria for clinical translation as a therapeutic neuromodulation target. Using resting-state fMRI in three independent repeated-imaging cohorts (healthy controls, early psychosis or chronic schizophrenia patients, minutes–months inter-scan intervals), CB-VTA FC was always stable and individual-specific (stability *r*=0.52–0.69; differential identifiability Δ*r*=0.21–0.35; all *p*<10⁻ L). In paravermal cerebellar territories, it tracked apathy severity in two patient cohorts (early psychosis, Crus I/II: *n*=99; *r*LL=0.36, *p*=2.65·10⁻L; chronic schizophrenia, Lobules VIIB/VIIIA: *n*=87 scans [65 patients]; *t*LL=4.06, *p*=1.1·10⁻L). In a meta-analysis of 39 randomized controlled transcranial magnetic stimulation trials (*n*=1,624; 867 active), connectivity of neighboring areas to stimulation site predicted negative symptoms improvement. CB-VTA FC thus emerges as a stable, individual-specific, and symptom-related therapeutically relevant circuit, constituting a mechanism-informed precision neuromodulation target in schizophrenia, ready for prospective clinical trials.

**One-sentence summary:** Cerebellum-ventral tegmental area functional connectivity is stable over time, differs across subjects, and its intensity is associated with the severity of apathy in schizophrenia.

## INTRODUCTION

Apathy is a prevalent and functionally debilitating symptom across neuropsychiatric disorders (1), with particularly high rates observed throughout all stages of schizophrenia spectrum illness (2). Characterized by reductions in motivation, pleasure and goal-directed behavior, apathy impacts real-world functioning across critical domains including housing stability, interpersonal relationships, educational attainment, and vocational performance (3). The socio-economic burden associated with apathy-related functional impairment is substantial, yet no approved pharmacological treatments exist, largely due to insufficient mechanistic understanding of the underlying neural circuitry (4).

Current neurobiological models of apathy in schizophrenia have predominantly focused on dysfunction within dopaminergic circuits supporting reward processing and motivational drive (5), which are implicated in the multidimensional construct of apathy (1). These circuits converge on the ventral tegmental area (VTA) and its projections to striatal and prefrontal regions (6). The deep anatomical location of these subcortical structures makes direct therapeutic intervention using non-invasive neuromodulation techniques challenging, prompting investigations of more accessible neural systems that might serve as surrogate targets for modulating these dopaminergic networks.

The cerebellum (CB) is a compelling candidate for such intervention, given its anatomical location and established connectivity with reward and motivational networks, in particular the VTA (7–11). In rodents, circuit-specific manipulation of cerebellar projections to the VTA bidirectionally controls motivated behavior: optogenetic activation of this pathway engages reward signaling and supports social preference, whereas silencing it abolishes social preference (9); chemogenetic activation and inhibition similarly modulate stress- and depression-related phenotypes (10). In humans, probabilistic tractography has evidenced mono- and polysynaptic cerebellum–VTA tracts originating predominantly in paravermal cerebellar regions, whose microstructural integrity correlates with self-reported socio-affective traits relevant to negative symptoms (11). Importantly, this circuit appears to hold pathophysiological relevance at the group level given that more frequent anti-coactivation of these same paravermal regions with the VTA was recently associated with reduced apathy severity in patients with schizophrenia (12), providing functional anchoring for the structural findings. From a therapeutic perspective, cerebellar stimulation through transcranial magnetic stimulation (TMS) effectively lowers negative symptoms of schizophrenia (13,14), and the cerebellar territories whose stimulation predicts symptom improvement overlap with the paravermal regions implicated in apathy (15), positioning the cerebellum as both mechanistically relevant to apathy *and* therapeutically accessible via non-invasive neuromodulation. Beyond group-level findings, prior dense-sampling work has demonstrated that cerebellar functional organization is substantially individual-specific in healthy adults (16), suggesting that personalized targeting via TMS may be warranted.

However, a systematic investigation of CB-VTA connectivity as a personalized neuromodulation target for apathy is currently lacking. Specifically, four critical gaps must be addressed (17):

*Stability*: Is CB-VTA functional connectivity (FC) reliable within individuals across time and across health and disease, a prerequisite for targeting?

*Individuality*: Do connectivity patterns vary sufficiently between individuals to enable personalization?

*Symptom-FC relationship*: Do CB-VTA FC patterns consistently associate with apathy severity across independent patient cohorts?

*Therapeutic relevance*: Does cerebellar FC relate to real-world treatment response?

Here, we address these gaps through a multi-dataset approach spanning healthy individuals and patients across the psychosis spectrum (**Figure 1**). We quantified the stability and differential identifiability of CB-VTA FC patterns using the Human Connectome Project Young Adults (HCP-YA, Dataset 1; 18) and Human Connectome Project Early Psychosis (HCP-EP, Dataset 2; 19) data, supplemented by a longitudinal clinical cohort of patients with chronic schizophrenia with age- and gender-matched controls (Dataset 3, scans acquired at baseline and at three-month follow-up). To examine the symptom-FC relationship, we tested associations between CB-VTA connectivity and apathy severity in patients. Finally, we leveraged meta-analytical data (15) and correlated normative functional connectivity derived from TMS sites in independent randomized clinical trials with the observed change in negative symptoms, to determine whether cerebellar territories identified in our direct analyses are also related to therapeutic response

**Figure 1:**
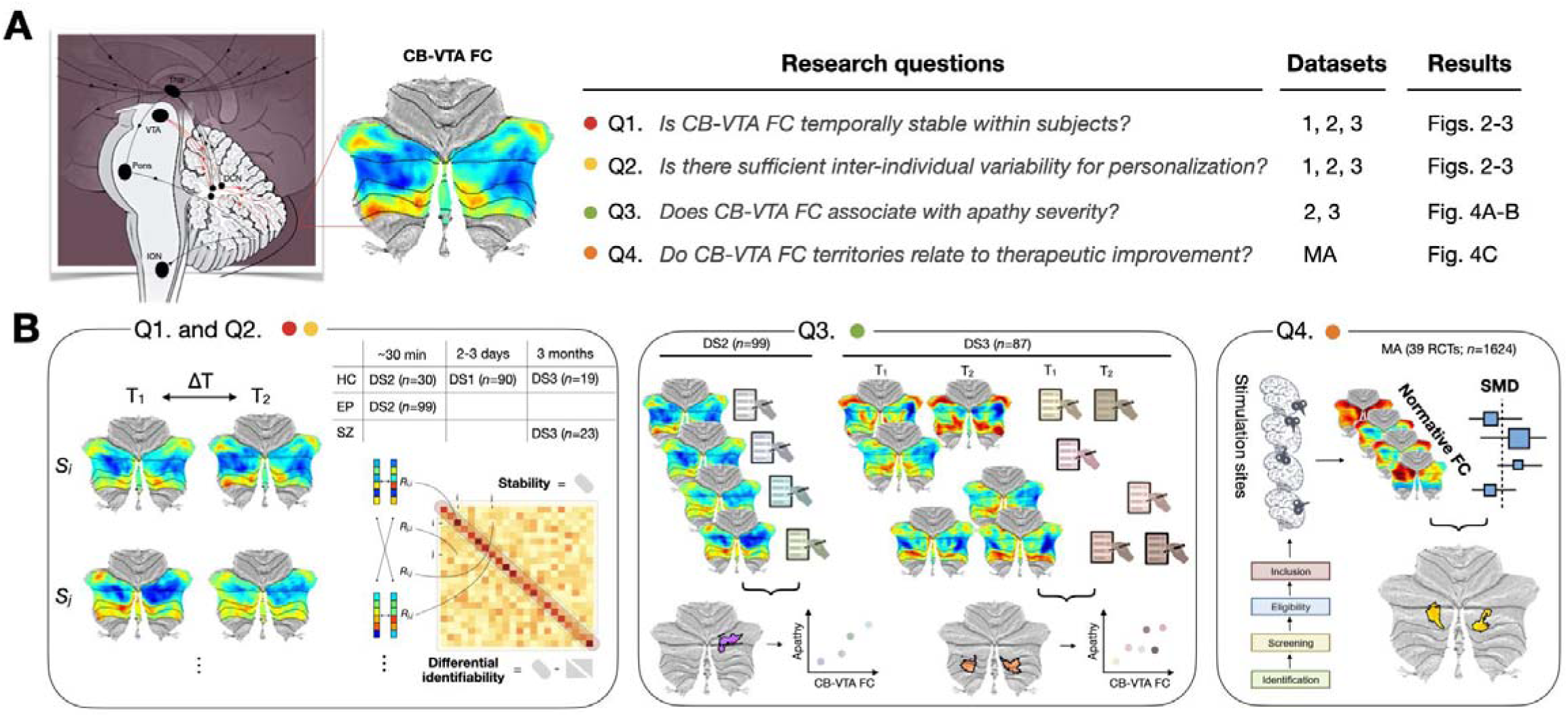
Core questions addressed in the study and leveraged datasets. **(A)** Schematic illustration of key brain nuclei involved in cerebellar signaling, with connections captured through cerebellar-ventral tegmental functional connectivity (CB-VTA FC) labeled in red, and connectivity of the VTA to other mesolimbic areas omitted for simplicity. The research questions addressed in this work are laid down alongside associated datasets and figures. **(B)** Schematic overview of employed methodology in terms of answered research questions. *Abbreviations:* VTA: ventral tegmental area, DCN: deep cerebellar nuclei, ION: inferior olivary nucleus, Thal: thalamus, CB-VTA FC: cerebellum-ventral tegmental area functional connectivity, MA: meta-analysis, DS: dataset, RCT: randomized controlled trial, SMD: standardized mean difference.

We hypothesized that CB-VTA FC would demonstrate significant within-subject stability and differential identifiability to support personalized targeting, while showing consistent relationships with apathy severity and treatment response across multiple independent datasets.

## RESULTS

### CB-VTA FC mapping

To determine whether CB-VTA FC is a measurable and reliable feature at the individual level, we extracted voxelwise cerebellar time courses and estimated VTA activity using a *seed map* approach (20,21). To do so, the activity time courses of whole-brain (sub)cortical voxels (excluding the cerebellum target region, shown in **Supplementary Figure 1A**) were weighted by their normative connectivity to the VTA (depicted in **Supplementary Figure 1B**), and averaged to yield a VTA reference signal. Consistent with past cortico-cortical FC findings (18), this method provided a higher signal-to-noise ratio than *seed-only* averaging (*i.e.*, computing VTA activity as the mere average of the activity time courses of VTA voxels), and captured subject-specific CB-VTA features with relatively short scan durations (see **Supplementary Methods – Constructing the time course of VTA activity** and **Supplementary Figure 2** for details).

### CB-VTA FC remains stable and individualized over hours to months across health and disease

We first assessed longitudinal stability of CB-VTA FC, a prerequisite for reliable clinical targeting.

In healthy adults from Dataset 1 (*n*=90 subjects), connectivity maps were highly consistent across sessions separated by several days, preserving individual fingerprints while differing markedly between subjects (**Figure 2A**), as shown by significant stability (*r*=0.69, 95% CI 0.67–0.71, *p*<10LL) and differential identifiability (Δ*r*=0.28, 95% CI 0.27–0.29, *p*<10LL). Similar observations could be made for the healthy controls from Dataset 2 (*n*=30 subjects, inter-scan interval: ∼30 min, **Supplementary Figure 3**).

**Figure 2:**
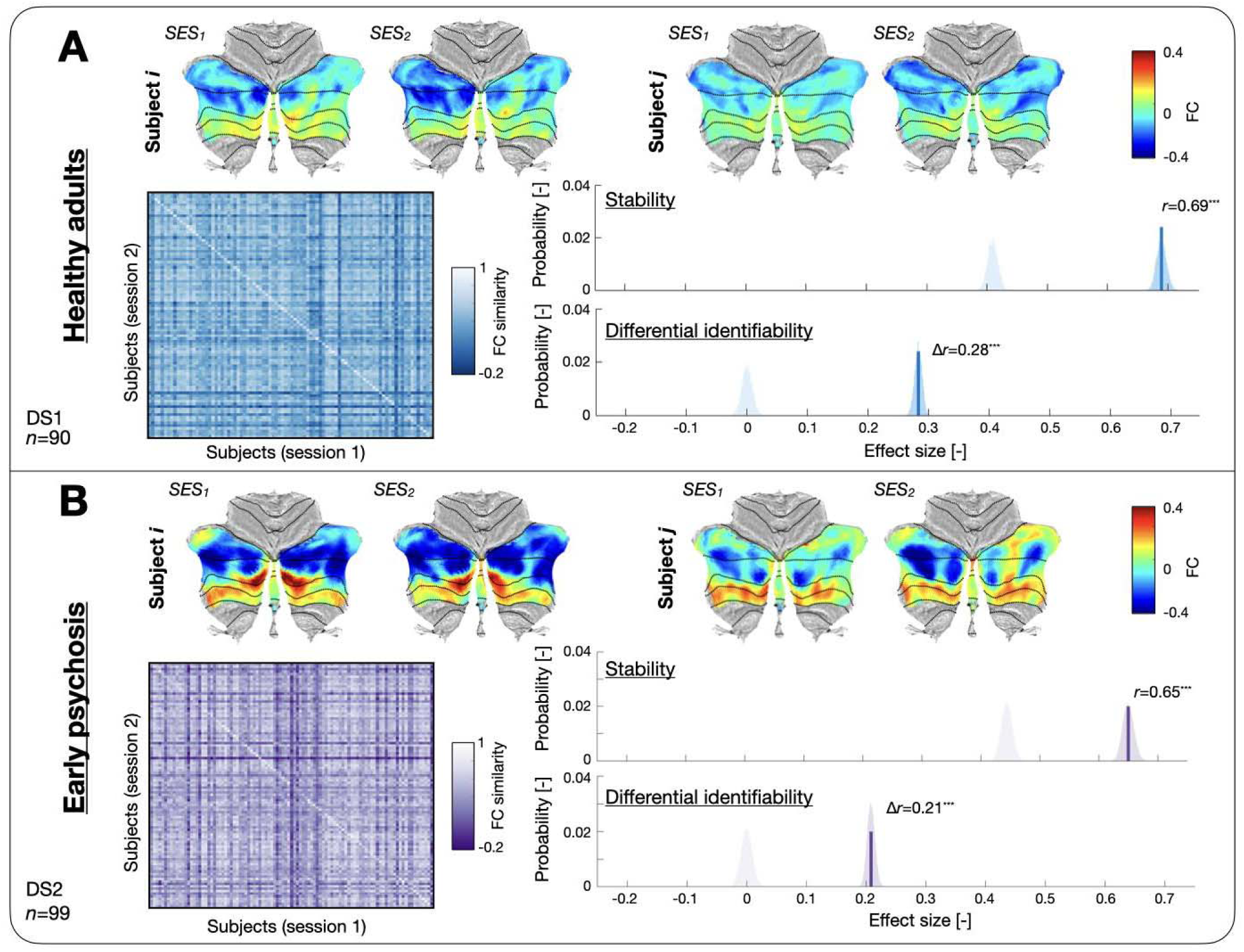
Cerebellum-ventral tegmental functional connectivity maps are stable and subject-specific in healthy subjects and early psychosis patients. **(A)** (*Top*) FC maps obtained for both resting-state sessions of two indicative subjects from Dataset 1. (*Bottom left*) Similarity between FC maps at the population level (*n*=90 young adults). Diagonal/off-diagonal elements reflect cross-session similarity values within or across subjects, respectively. (*Bottom right*) Stability (mean of diagonal similarity elements) was significant (top panel, *r*=0.69, *p*<10^-5^, light/dark blue null distribution/bootstrapped confidence interval), and so was differential identifiability (difference between mean [off-]diagonal similarity values; Δ*r=*0.28, *p*<10^-5^). **(B)** Similar displays for 99 early psychosis patients. Stability (mean of diagonal similarity elements) was significant (top panel, *r*=0.65, *p*<10^-5^, light/dark purple null distribution/bootstrapped confidence interval), and so was differential identifiability (difference between mean [off-]diagonal similarity values; bottom panel, Δ*r=*0.21, *p*<10^-5^). *Abbreviations:* SES: session, FC: functional connectivity.

Patients with early psychosis from Dataset 2 (*n*=99 subjects) showed reproducible and individualized maps at a timescale of around half an hour (**Figure 2B**; stability: *r*=0.65, 95% CI 0.63–0.67, *p*<10LL; differential identifiability: Δ*r*=0.21, 95% CI 0.2–0.22, *p*<10LL). Even over three months in Dataset 3, stability and differential identifiability remained significant in patients and age- and gender- matched healthy controls (HCs, *n*=19 subjects: *r*=0.52, 95% CI 0.47–0.57; Δ*r*=0.27, 95% CI 0.22–0.31; SZ patients, *n*=23 subjects: *r*=0.54, 95% CI 0.5–0.59, Δ*r*=0.35, 95% CI 0.32–0.38; all *p*-values<10LL; **Figure 3**). Thus, across healthy, early psychosis and chronic schizophrenia cohorts, CB-VTA FC proved significantly reproducible and individualized over timescales of minutes to days and months. Furthermore, neither stability nor differential identifiability was consistently associated with age, sex, or race across cohorts (see **Supplementary Results – Association of stability and differential identifiability with demographic and clinical measures**). Stability and differential identifiability results are summarized, for all datasets, in **Table 1**.

**Figure 3:**
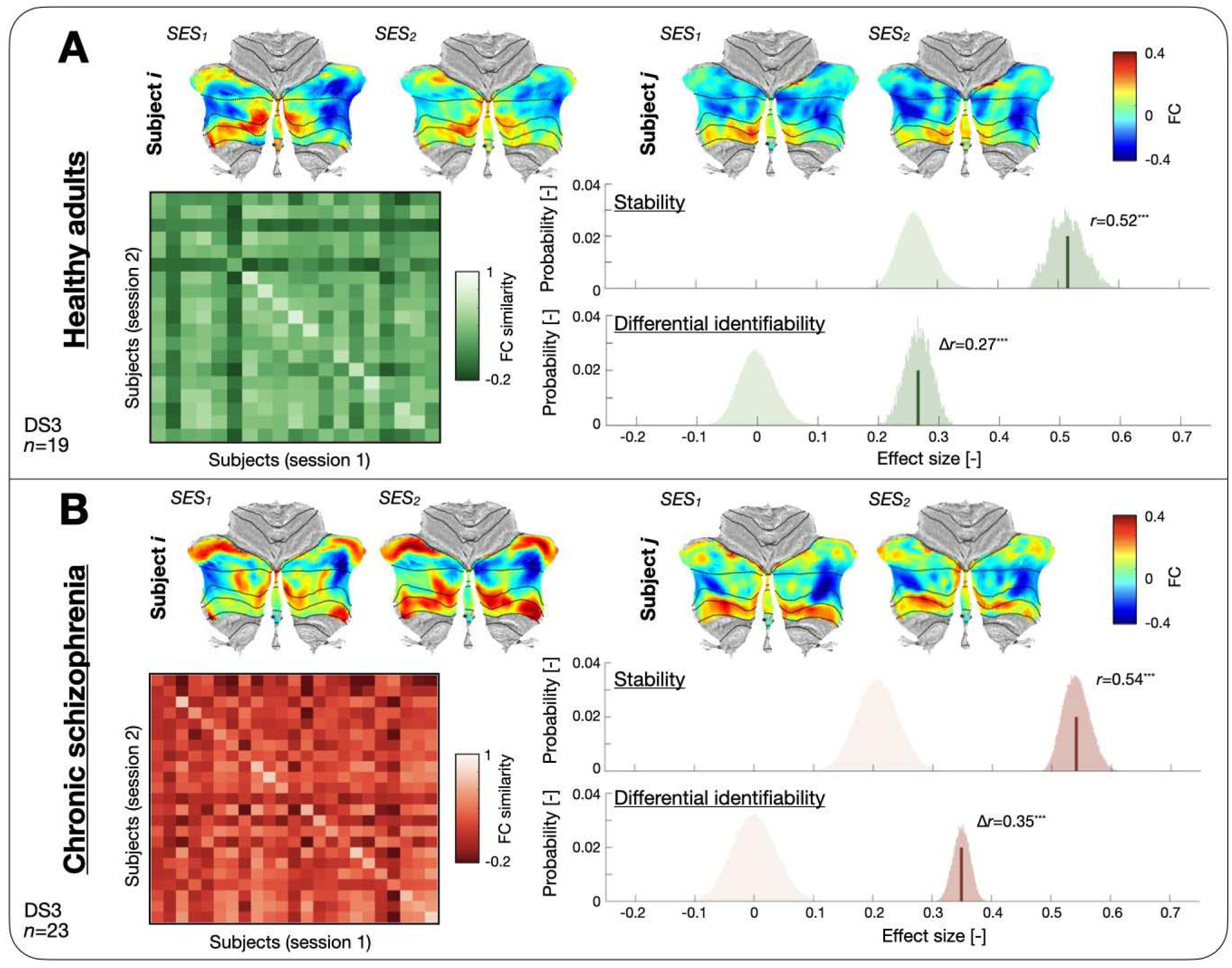
Cerebellum-ventral tegmental functional connectivity maps are stable and subject-specific over 3 months in patients with chronic schizophrenia and matched controls. **(A)** (*Top*) FC maps obtained for both resting-state sessions of two healthy controls from Dataset 3. (*Bottom left*) Similarity between FC maps at the population level for 19 healthy controls. Diagonal/off-diagonal elements reflect cross-session similarity values within or across subjects, respectively. (*Bottom right*) Stability (mean of diagonal similarity elements) was significant (top panel, *r*=0.52, *p*<10^-5^, light/dark green null distribution/bootstrapped confidence interval), and so was differential identifiability (difference between mean [off-]diagonal similarity values; Δ*r=*0.27, *p*<10^-5^). **(B)** Similar displays for 23 patients with chronic schizophrenia patients. Stability (mean of diagonal similarity elements) was significant (top panel, *r*=0.54, *p*<10^-5^, light/dark red null distribution/bootstrapped confidence interval), and so was differential identifiability (difference between mean [off-]diagonal similarity values; bottom panel, Δ*r=*0.35, *p*<10^-5^). *Abbreviations:* SES: session, FC: functional connectivity.

**Table 1:** Summary of stability and differential identifiability results across datasets. For all investigated datasets, stability and differential identifiability (DI) results are summarized, including descriptive statistics (mean and standard deviation across bootstrapping folds), 95% confidence intervals (CI) across bootstrapping folds, and *p*-values (inferred from null permutations). EP, early psychosis; SZ, schizophrenia; ISI, inter-scan interval.

| Dataset |  |  | Stability |  |  | DI |  |  |
| --- | --- | --- | --- | --- | --- | --- | --- | --- |
| Name | Group | ISI | mean ± std | 95% CI | *p*-value | mean ± std | 95% CI | *p*-value |
| DS1 (HCP-YA) | Healthy | Days | 0.69±0.008 | [0.67, 0.71] | <10<sup>-5</sup> | 0.28±0.006 | [0.27, 0.29] | <10<sup>-5</sup> |
| DS2 (HCP-EP) | Healthy | ~30 min | 0.68±0.01 | [0.66, 0.7] | <10<sup>-5</sup> | 0.24±0.01 | [0.21, 0.26] | <10<sup>-5</sup> |
| DS2 (HCP-EP) | EP | ~30 min | 0.65±0.009 | [0.63, 0.67] | <10<sup>-5</sup> | 0.21±0.007 | [0.2, 0.22] | <10<sup>-5</sup> |
| DS3 (local) | Healthy | ~3 months | 0.52±0.03 | [0.47, 0.57] | <10<sup>-5</sup> | 0.27±0.02 | [0.22, 0.31] | <10<sup>-5</sup> |
| DS3 (local) | Chronic SZ | ~3 months | 0.54±0.02 | [0.5, 0.59] | <10<sup>-5</sup> | 0.35±0.01 | [0.32, 0.38] | <10<sup>-5</sup> |

### Intensity of CB-VTA FC correlates with apathy

If CB-VTA FC is to serve as a personalized and mechanistically informed therapeutic target for apathy, it must show a meaningful association with apathy severity.

In early psychosis patients (Dataset 2, *n*=99 subjects; analyses adjusted for antipsychotic medication and psychosis subtype), stronger CB-VTA anticorrelation was significantly associated with less severe apathy in a cluster situated in the right paravermal cerebellum in Crus I/II regions (**Figure 4A**). CB-VTA FC was negative on average in this cluster (cluster C_1_: FC=-0.13±0.08, cluster extent = 215 voxels; *t-*statistics: min=2.63, max=3.38, mean [median]=2.88 [2.86]). Therefore, CB-VTA anticorrelation in Crus I/II appears to reflect lower apathy severity, whereas reduced anticorrelation relates to higher apathy, directly replicating prior reports (12).

**Figure 4:**
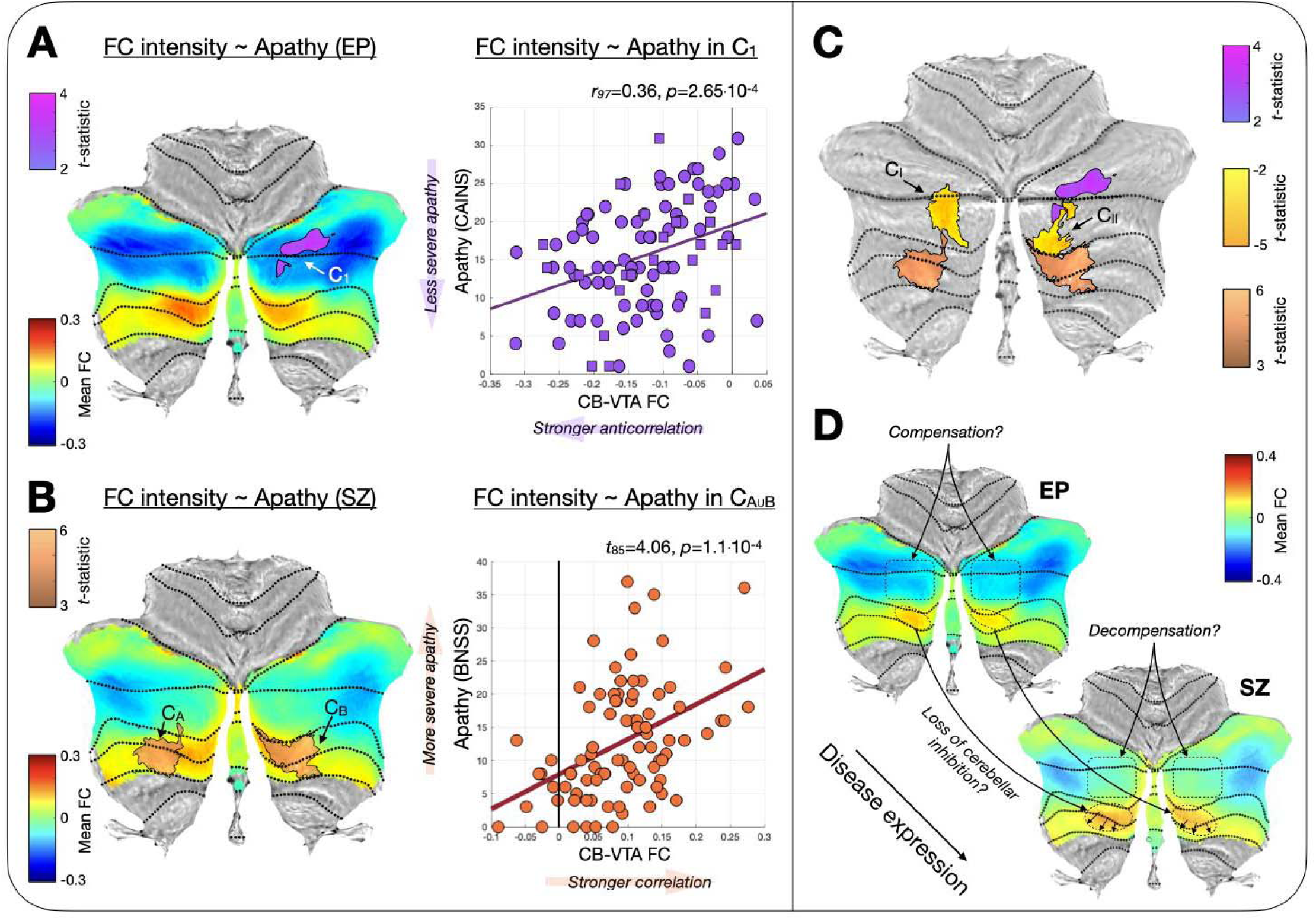
Cerebellum-ventral tegmental functional connectivity correlates with apathy in early psychosis and chronic schizophrenia patients. **(A)** (*Left*) Significant cluster (labeled C_1_, shown in purple and indicated by a white arrow) for the correlation between CB-VTA FC and apathy in early psychosis patients (Dataset 2, *n*=99 patients, corrected for antipsychotic dosage and psychosis subtype). Population-wise average CB-VTA FC is shown as an underlay. (*Right*) Association between C_1_ mean CB-VTA FC across the cluster and apathy (*r*=0.36, *p*=2.65·10^-4^). Squares/circles denote the data from patients with affective/non-affective psychosis, respectively. **(B)** (*Left*) Significant clusters (labeled C_A_ and C_B_, shown in copper color and indicated by black arrows) for the correlation between CB-VTA FC and apathy in chronic schizophrenia patients (Dataset 3, *n*=87 scans from 65 patients, corrected for antipsychotic dosage). Population-wise average CB-VTA FC is shown as an underlay. (*Right*) Association between C_AUB_ mean CB-VTA FC across the combined clusters and apathy (*t*_85_=4.06, *p*=1.1·10^-4^). **(C)** Clusters where cerebellar FC to the TMS site significantly correlated with SMD (*p*<0.05 FWE-corrected; C_I_ and C_II_, displayed in yellow and labeled with black arrows), shown alongside the clusters found in **(A)** and **(B)**. **(D)** Conceptual proposal for how CB-VTA FC may evolve from early psychosis to chronic schizophrenia, including a spatial spread of coupling within Lobules VIIB/VIIIA denoting the loss of cerebellar inhibition on the VTA, and a compensatory anticoupling in Crus I/II that remains effective in early psychosis, but then dissipates in chronic schizophrenia (decompensation). *Abbreviations:* EP, early psychosis, FC: functional connectivity, C_1_: cluster 1, CAINS: Clinical Assessment Interview for Negative Symptoms, CB-VTA FC: cerebellum-ventral tegmental area functional connectivity, SZ: chronic schizophrenia, C_AUB_: union of clusters A and B, BNSS: Brief Negative Symptom Scale, C_I_: cluster I, C_II_: cluster II.

If anticorrelation indicates preserved motivational circuitry, then it is reasonable to hypothesize that its loss (*e.g.*, as illness progresses) would relate to more severe apathy. In other words, positive coupling would result in worse apathy. In line with this, in patients with chronic schizophrenia (Dataset 3, *n*=87 scans from 65 patients; analyses adjusted for antipsychotic medication), more severe apathy was related to more positive correlation of the CB with the VTA (**Figure 4B**) in two bilateral paravermal clusters in Lobules VIIB/VIIIA. In these clusters (C_A_ and C_B_), CB-VTA FC was positive on average (FC=0.1±0.07 and 0.08±0.09, respectively). C_A_ comprised 164 voxels (*t-*statistics: min=3.28, max=5.62, mean [median]=3.86 [3.66]), and C_B_ comprised 158 voxels (3.27, 5.0, 3.77 [3.73]). Similar observations could be made when individually analyzing each cluster (**Supplementary Figure 4**).

### Cerebellar-VTA FC indicates clinical recovery upon TMS

Finally, if the paravermal clusters that we identified empirically are mechanistically relevant to apathy, their FC should relate to real-world treatment response. We therefore tested whether stimulation sites from 39 randomized controlled trials of TMS in schizophrenia are associated with improvement in negative symptoms (defined as mean change in effect size from active versus sham stimulation), by mapping each site to normative functional connectivity and testing for associations with symptom change.

Functional connectivity of stimulation sites showing greater therapeutic benefit converged on bilateral Crus I/II cerebellar regions (**Figure 4C**, clusters C_I_ and C_II_). C_I_ and C_II_ comprised 103 (*t*-statistics: min=-3.17, max=-2.06, mean [median]=-2.46 [-2.4]) and 98 (−3.18, −2.06, −2.39 [-2.33]) voxels, respectively. In both clusters, more negative-valued functional connectivity to the stimulation site was linked to greater improvement in symptoms. Convergence zones were directly adjacent to C_1_, C_A_ and C_B_.

## DISCUSSION

In this work, we provided converging evidence from multiple datasets that CB-VTA FC has potential to become a clinically relevant target network for apathy in schizophrenia. Through systematic evaluation across four critical translational criteria (17) – stability, differential identifiability, symptom-circuit relationships, and therapeutic relevance – our multi-dataset approach demonstrates that CB-VTA connectivity patterns are robust, personalized, and mechanistically meaningful to guide prospective precision neuromodulation interventions.

CB-VTA FC demonstrated remarkable stability across timescales from minutes to months (*r*=0.52–0.69, all *p*-values<10LL), spanning healthy individuals, early psychosis patients, and chronic schizophrenia (**Figures 2-3**). This temporal consistency is essential for reliable clinical targeting, as connectivity patterns must remain stable between baseline assessment and therapeutic intervention. Equally critical is inter-individual variability to justify personalized targeting. Our differential identifiability metrics (Δ*r*=0.21–0.35, all *p*-values<10LL) indicate that while connectivity patterns are stable within individuals, they vary meaningfully between subjects, consistent with previous findings using dense sampling (16) – providing the foundation for personalized circuit-guided interventions. This balance between stability and individuality positions CB-VTA connectivity as a candidate circuit for precision psychiatry approaches.

A novel contribution of this study is the demonstration of coherent symptom–circuit relationships that appear to evolve across illness stages in independent cohorts, potentially delineating the pathophysiological trajectory of apathy in schizophrenia. In early psychosis, greater apathy correlated with reduced anticorrelation between the cerebellar paravermis (*i.e.*, medial cerebellum juxtaposed with the vermis; Crus I/II) and the VTA (**Figure 4A**). This pattern could indicate a partial weakening of cerebellar inhibitory control, while the fundamental antagonistic relationship between the regions remained intact. The persistence of anticorrelation suggests that cerebello–VTA gating remains functionally active, although circuit disruption may already have begun, as reflected by emerging pockets of positive coupling already present within paravermal regions (Lobules VIIB/VIIIA bilaterally). In chronic schizophrenia, a loss of anticorrelation was observed in Crus I/II, and greater apathy severity was associated with stronger positive correlation in Lobules VIIB/VIIIA bilaterally (**Figure 4B**). This shift from anticorrelation to frank positive correlation in spatially contiguous regions suggests progressive disinhibition of cerebellar outputs, consistent with circuit-level decompensation. Importantly, these relationships were independent of antipsychotic dosage and consistently involved paravermal cerebellar regions, in line with previous findings (12). These CB-VTA connectivity changes may parallel previously reported cerebello–thalamo–cortical hyperconnectivity, a robust and state-independent neural signature for psychosis prediction (22). In particular, increased connectivity within cerebello–thalamo–cortical circuits has been observed in individuals who later convert to psychosis compared with non-converters and predicts long-term outcome (23). Similarly, cerebello-cortical hyperconnectivity has been found in schizophrenia between Crus I, Crus II and cortical regions, in particular within the salience (ventral attention) network (24). Together, these findings suggest that the observed CB–VTA (hyper)coupling could reflect a broader trajectory of maladaptive cerebellar circuit reorganization across illness stages.

Converging neurochemical, cellular and circuit-level evidence supports the interpretation of CB–VTA coupling abnormalities stemming from cerebellar inhibition reduction. Postmortem studies reveal the altered expression of genes involved in vesicular transport, glutamatergic (as mediated by NMDA receptors) and GABAergic transmission, as well as calcium buffering in the cerebellar cortex of patients with schizophrenia (25–27). Purkinje cells have particularly high metabolic activity and are most vulnerable to homeostatic disruption (28). First-episode psychosis is associated with elevated cerebellar and striatal glutamate, but antipsychotic treatment only normalized the latter but not the former (29), suggesting that cerebellar glutamatergic dysfunction may reflect an upstream or independent excitatory imbalance. In line with these findings, strikingly, the (Glutamate + Glutamine)/GABA ratio correlates with symptom severity and normalizes after cerebellar TMS (30). Collectively, the alteration of cerebellar modulatory excitatory and inhibitory circuitries in schizophrenia, concords with evidence pointing to E/I imbalance as a ‘master mechanism’ of schizophrenia (31). According to the model proposed by Howes *et al.* (31), dysregulated cortical and subcortical inputs drive midbrain dopaminergic hyperactivity: here, we provide convergent proof to extend this framework by identifying cerebellar disinhibition as a key upstream contributor to this process. As schematically illustrated in **Figure 4D**, we propose that cerebellar inhibition is gradually lost from early psychosis to chronic schizophrenia, as reflected by the expansion of pockets of CB-VTA correlation in Lobules VIIB/VIIIA. Crus I/II anticorrelation with the VTA may be the reflection of compensatory mechanisms, which remain effective in early psychosis, but gradually fade upon progression of the disease (decompensation).

Meta-analytic evidence indicates that TMS sites whose normative connectivity overlaps with VTA-linked cerebellar territories yield greater improvement in negative symptoms, underscoring the translational potential of this circuit. Cerebellar projections can modulate distant forebrain circuits, bridging rodent and human evidence (32–34). In rodents, cerebellar output regulates distant (*i.e.*, prefrontal) dopamine release via the VTA and restores schizophrenia-related frontal dysfunction (32,33), while in humans, cerebellar stimulation engages large-scale cortical networks in a topographically specific manner (34). The cerebellum thus emerges as a particularly promising neuromodulation target given an established clinical safety record (35) and circuit-specific behavioral effects beyond stimulation site alone (36). Together, these data delineate a mechanistic pathway from cellular damage to system-level cerebellar dysfunction and define a rational target for circuit-based interventions in schizophrenia.

### Strengths and limitations

This is the first study to demonstrate replication of CB–VTA connectivity–symptom associations across three independent datasets spanning health, early psychosis, and chronic schizophrenia. Notably, replication held despite substantial differences in cohort characteristics – including illness stage, clinical phenotyping instruments (CAINS versus BNSS), scanner platforms, and acquisition protocols – strengthening confidence that the identified paravermal substrate reflects a genuine pathophysiological mechanism of apathy in schizophrenia. Further strengths include the integration of multimodal approaches linking neuroimaging, symptom associations, and meta-analytic evidence. Importantly, the cerebellar territories implicated by our functional analyses converge with regions independently identified in the structural connectivity literature, including diffusion MRI tractography of cerebello-midbrain pathways (11), and with sites whose stimulation in randomized clinical trials yields therapeutic benefit (15). Convergence across structural, functional, and interventional modalities is rare and provides compelling evidence for the circuit’s mechanistic relevance. Our data-driven, voxelwise approach further supports the rigor of the identified cerebellar clusters and their robustness as candidate targets for circuit-guided intervention in a symptom domain that remains largely refractory to conventional pharmacological treatment. We also demonstrated reproducibility across samples and sufficient inter-individual variability to support personalized applications. By anchoring our results to randomized controlled trial data, we strengthen the case for clinical translation in a domain of psychiatry with substantial unmet therapeutic needs.

Limitations include the small size and low signal-to-noise ratio of the VTA, which may constrain spatial precision. To enhance reliability, the seed incorporated weighted contributions from neighboring subcortical regions strongly connected to the VTA (20,21); consequently, CB–VTA connectivity reflects coupling with the broader VTA-associated network. Further, our suggested timeline of CB-VTA FC changes with disease expression (**Figure 4D**) should only be taken as a conceptual proposal, as Datasets 2 and 3 were acquired with distinct protocols and could thus not be quantitatively contrasted. However, both the loss of Crus I/II anticorrelation and positive correlations of Lobules VIIB/VIIIA are also observed in SZ patients as compared to matched HCs in Dataset 3 (**Supplementary Figure 5**), fully aligning with our hypotheses and opening the perspective for future investigations. Another limitation is that, Neurosynth-derived cerebellar maps are biased by field-of-view truncation in primary studies and to the under-representation of cerebellar findings in the literature (37), and may therefore be conservative for posterior cerebellar territories. In this context, the meta-analytical convergence with our direct analyses under these conservative conditions arguably strengthens rather than weakens the inference. Finally, a consideration concerns the spatial granularity at which CB–VTA connectivity differs between individuals (a set of characteristic maps can be appreciated across cohorts in **Supplementary Figure 6**). Given the relatively conserved cerebellar macroanatomy, inter-individual variability may manifest at a spatial scale finer than the focality of conventional figure-of-eight TMS coils. However, even small spatial differences in TMS targeting appear to carry meaningful functional consequences: compelling work has shown that identical TMS patterns applied to nearby cortical sites subserving distinct brain circuitry elicit opposite local metabolic effects (36). Future work should quantify the spatial granularity of inter-individual CB–VTA variability against the envelope focality of candidate stimulation modalities, including high-focality TMS coil designs, neuronavigation-guided electric-field modeling, and complementary approaches such as transcranial focused ultrasound, to refine precision targeting strategies.

## MATERIALS AND METHODS

### Study design

Specific details regarding the determination of final sample sizes (available data, determination of required sizes for end measures at hand and removal of outliers) and the inclusion/exclusion criteria across analyzed datasets can be found in the **Overview of examined datasets**, **Data collection background** and **Determination of final sample sizes** subsections below.

As further detailed in the **Overview of examined datasets** subsection below, our end measures were (1) CB-VTA stability, (2) CB-VTA differential identifiability and (3) associations between CB-VTA FC and apathy. Our research objectives were to (1) assess whether stability and differential identifiability of CB-VTA FC were significant in healthy individuals and patients suffering from schizophrenia at various stages of disease expression, and (2) unravel cerebellar territories with significant associations between CB-VTA FC and symptoms of apathy in patients across the course of the disease. To achieve these goals, we analyzed a combination of healthy individuals, patients with early psychosis, and patients with chronic schizophrenia.

### Overview of examined datasets

Our analyses included unrelated healthy adults from the Human Connectome Project Young Adults (HCP-YA) dataset (*n*=90, referred to as Dataset 1 from here onwards), patients with early psychosis from the HCP Early Psychosis (HCP-EP) dataset (*n*=99, Dataset 2), and patients with chronic schizophrenia (SZ, *n*=23) and matched healthy controls (HCs, *n*=19) whose data was acquired locally (Dataset 3).

Dataset 1 was used to quantify CB-VTA FC stability and differential identifiability at the time scale of a few days in healthy subjects (**Figure 1B**, Q1. and Q2.; **Figure 2A**). Dataset 2 was used to quantify CB-VTA FC stability and differential identifiability at the time scale of half an hour in patients with early psychosis (**Figure 1B**, Q1. and Q2.; **Figure 2B**), and to link CB-VTA FC to negative symptoms of apathy (**Figure 1B**, Q3.; **Figure 4A**). The Clinical Assessment Interview for Negative Symptoms (CAINS), a thirteen-item scale endorsed by the NIMH’s Consensus Development Conference on Negative Symptoms (38), was used to quantify negative symptoms. Apathy was calculated as the sum of items 1-9, *Motivation and Pleasure* (MAP) subscale.

Dataset 3 was used to quantify CB-VTA FC stability and differential identifiability at the time scale of three months in healthy controls and patients with chronic schizophrenia (**Figure 1B**, Q1. and Q2.; **Figure 3**). After quality control, and including subjects with one or two usable scans, 87 scans from SZ patients were retained to link CB-VTA FC to negative symptoms of apathy (**Figure 1B**, Q3.; **Figure 4B**). To quantify negative symptoms, we used the Brief Negative Symptoms Scale (BNSS; 39), a semi-structured interview conducted by trained researchers/clinicians.

The BNSS quantitatively measures negative symptoms across five individual symptoms (anhedonia, asociality, avolition, flat affect, alogia). BNSS items are scored on a seven-point severity scale, higher scores indicating more severe impairment. The apathy subscale score was calculated as the sum of the anhedonia, asociality and avolition items.

### Data collection background

#### Dataset 1 (Human Connectome Project Young Adults, HCP-YA)

Most participants were born in Missouri, and recruiting was performed to ensure a fair representation of the ethnic and racial composition of the United States of America. Included subjects were representative of the population at large, to capture the variability present across healthy individuals in behavioral, ethnic and socioeconomic terms. Smokers, overweight individuals, or people with a history of heavy drinking or recreational drug use without having experienced severe symptoms, were included. Subjects were drawn from a population of adult twins and their non-twin siblings. Participants were in the age range of 22–35 years. Candidates with siblings suffering from severe neurodevelopmental, neuropsychiatric or neurologic disorders were excluded. A detailed list of inclusion and exclusion criteria can be found in **Supplementary Table 1**.

HCP subjects underwent a detailed cognitive assessment revolving around both task-based paradigms performed in the scanner, and in-depth out-of-scanner assessments (see

https://www.humanconnectome.org/storage/app/media/documentation/s1200/HCP_S1200_Release_Reference_Manual.pdf for details). In this work, we did not perform any analyses related to these scores.

Participants all underwent a practice session in a mock scanner before any actual scan, including feedback on head movement and training to minimize it. Resting-state scans were acquired over the course of two separate days, within a two- to three-day period according to feasibility. Two resting-state scans were acquired on Day 1, and two others on Day 2, for a total 57.6 min of data (4 scans long of 14.4 min each at TR=0.72 s) per subject. Further details regarding data collection can be found in (18).

#### Dataset 2 (Human Connectome Project Early Psychosis, HCP-EP)

The HCP-EP dataset is a follow-up on the HCP initiative, aimed at publicly sharing clinical, cognitive, behavioral, functional and structural neuroimaging data to characterize the pathological substrates across the psychosis spectrum within the first 5 years following illness onset, in individuals with affective and non-affective psychosis.

Participants (16-35 years old) were recruited at 3 sites in Massachusetts and one site in Indiana, with data collection matched across sites, from local early psychosis programs. Patients had to meet either of the following inclusion diagnostic criteria:

1. DSM-V diagnosis of schizophrenia, schizophreniform disorder, schizoaffective disorder, psychosis not otherwise specified, delusional disorder, or brief psychotic disorder with onset within the past five years prior to study entry (non-affective psychosis)
2. DSM-V diagnosis of major depression with psychosis (single and recurrent episodes) or bipolar disorder with psychosis (including most recent episode depressed and manic types) with onset within five years prior to study entry (affective psychosis).

Notorious exclusion criteria (see **Supplementary Table 2** for an exhaustive list of inclusion and exclusion criteria) included substance-induced psychosis or psychotic disorder due to a medical condition, subjects with an active medical condition that affects brain or cognitive functioning, current severe substance use disorder in the past 90 days (excluding caffeine and nicotine), and electroconvulsive therapy treatment in the past year.

HCP-EP subjects underwent a detailed clinical and cognitive assessment (see https://www.humanconnectome.org/storage/app/media/documentation/HCP-EP1.1/HCP-EP_Release_1.1_Manual.pdf for details). For all patients, lifetime antipsychotic medication dosages were calculated as chlorpromazine (CPZ) equivalents using the Gardner approach (40). Further details about data collection and clinical evaluation can be found in (19).

Study procedures (including resting-state scanning and cognitive/clinical assessments) took place on the same day. Two pairs of resting-state scans were acquired with a delay of approximately 24.89 min, yielding a total of 21.87 min of data per subject (5.47 min per scan at TR=0.8 s).

##### Dataset 3

All participants provided informed consent under protocols approved by the Geneva Ethics Committee (CCER. BASEC IDs: 2017-01765, 2020-02169). We recruited clinically stable patients with chronic schizophrenia (*i.e.*, no changes in medication and no hospitalizations in the last 3 months) with confirmed schizophrenia spectrum diagnosis via the Mini International Neuropsychiatric Interview (41), and matched healthy controls.

Given our focus in obtaining mechanistic insights and in accordance with recent recommendations (4), we excluded patients with secondary sources of negative symptoms (*e.g.*, state of active psychosis, extrapyramidal effects, medication, depression, sedation; see below for exclusion criteria). Patients were recruited from community outpatient centers in the University Hospital of Geneva, Switzerland.

Patients were not included if they presented other Axis-I disorders, in particular comorbid major depressive episodes, florid psychotic symptoms [Positive and Negative Syndrome Scale, any positive item >4 (42)], significant extrapyramidal side effects [any St. Hans Rating Scale item >3 (43)], use of benzodiazepines or lorazepam equivalents of more than 1 mg, or active use of substances (including opiate substitution for medical purposes).

Healthy controls were included only if they did not present any Axis-I disorder, a family history of psychotic disorders or psychotropic drug use.

All Dataset 3 individuals (HCs and SZ patients) underwent a detailed psychopathological and cognitive assessment at baseline (T_1_) and at three-month follow-ups (T_2_). In addition to the BNSS, we also employed the Positive and Negative Syndrome Scale [PANSS; (42)], the Calgary Depression Scale for Schizophrenia [CDS; (44)], and the Personal and Social Performance Scale [PSP; (45)]. Cognitive function was assessed with the Brief Assessment of Cognition in Schizophrenia (BACS) battery (46), and extrapyramidal syndrome was assessed with the St. Hans Rating Scale (43). Antipsychotic medication was recorded in risperidone equivalents.

Resting-state functional imaging was also performed both at T_1_ and T_2_, acquiring 9.8 min of data each time at TR=1 s.

From 427 individuals screened, we recruited 146 (90 SZ patients and 56 HCs) at T_1_, 65 of which (36 SZ patients and 29 HCs) were also examined at T_2_. A subset of the cohort included in this study was drawn from a longitudinal study (BASEC ID: 2017-01765).

Note that part of this dataset has already been analyzed in (12), but using co-activation pattern (CAP) analysis, a distinct time-resolved methodology (47), while here we entirely focus on static FC estimates.

#### Data acquisition

##### Datasets 1 and 2

Acquisition details largely resembled each other between Datasets 1 and 2, with a few specific differences as highlighted below.

All Dataset 1 (HCP-YA) subjects underwent structural MRI, diffusion MRI, as well as task-based and resting-state functional MRI. They were scanned on a customized Siemens 3T Connectome Skyra at Washington University, using a standard 32-channel Siemens receive head coil. The resting-state data was acquired in 4 fifteen-minute runs. Phase encoding was performed in the right-to-left, then left-to-right direction for two sessions, and in the reverse order for the remaining two, partly to enable the compensation of dropout signal by combining two scans with opposing phase encoding direction.

Acquisitions were based on the blood oxygenation level-dependent (BOLD) contrast, using gradient echo echo-planar imaging with a multiband acceleration factor of 8. The acquired data had the following properties: field of view = 208 mm × 180 mm × 144 mm (anterior-posterior × phase-encoding × inferior-superior), 72 slices (*i.e.*, 9 groups of 8 simultaneously acquired slices), resolution = 2 mm isotropic, TR = 720 ms, 1200 acquired volumes (14 min 24 sec).

In a darkened room environment, the subjects were asked to lie with eyes open and to fixate a white cross while remaining relaxed, thinking of nothing in particular and not falling asleep. More details on acquisition can be found in (48) for how HCP parameters optimize the trade-off between spatial and temporal resolutions, and in (49) for the specifics of resting-state acquisitions in the HCP.

The HCP-EP (Dataset 2) data was acquired on 4 Siemens 3T Magnetom Prisma scanners at Beth Israel Deaconess Medical Center, McLean Hospital, Massachusetts General Hospital, and Indiana University. The McLean data used a 64-channel head and neck coil, with neck channels turned off, while at other sites, a 32-channel head coil was used. Acquisitions followed similar procedures as in the HCP-YA case. Participants fixated a black cross on a light background, and the TR was set to 800 ms. Each resting-state session lasted 5 minutes and 28 seconds (410 volumes).

##### Dataset 3

For Dataset 3, MRI data were acquired at the Campus Biotech (Geneva, Switzerland) using a Siemens 3T Magnetom Prisma scanner and a standard 64-channel head coil. High-resolution structural images were captured utilizing a T1-weighted sequence with specified parameters (voxel size = 1.0 × 1.0 × 1.0 mm^3^, repetition time = 2200 ms, echo time = 2.96 ms, field of view read = 256 mm, flip angle = 9°, slice thickness = 1.0 mm, phase encoding: anterior-to-posterior, no fat suppression) to ensure optimal volumetric resolution. Resting-state functional MRI scans were acquired with the following parameters: 589 frames (9.8 minutes), field of view read = 224 mm, voxel size = 2.0 mm isotropic, 66 transversal slices, slice thickness = 2.0 mm, TR = 1000 ms, echo time = 32.0 ms, flip angle = 50°, phase encoding: anterior-to-posterior, acceleration mode with factor for parallel imaging = 6. Full details can be found in (12).

#### Data preprocessing

##### Spatial preprocessing

Datasets 1 and 2 underwent standard HCP preprocessing including gradient nonlinearity correction, motion correction via single-band reference registration, BL distortion correction, and structural registration. All transforms were concatenated and applied in a single resampling to 2 mm MNI space.

Dataset 3 was preprocessed using SPM12 https://www.fil.ion.ucl.ac.uk/spm/software/spm12/ with standard realignment, coregistration, segmentation to extract tissue probability maps, and nonlinear normalization to MNI space. Processing quality was visually inspected at each step.

##### Preprocessing of voxelwise time courses

For Datasets 1 and 2, global intensity normalization by a single scaling factor was applied, and non-brain voxels were masked out, followed by the application of minimal high-pass filtering (cutoff: 0.0005 Hz). Next, Independent Component Analysis was run with MELODIC (50) with automatic dimensionality estimation, limiting the dimension to 250. Resulting components were fed into FMRIB’s ICA-based X-noiseifier (FIX, 51), and components classified as “bad” (*e.g.*, representative of head motion or cardiac pulsation) were removed from the data. The HCP-YA and HCP EP data were provided at this stage as “FIX-denoised” data for download. From there, identical further steps were applied to all the analyzed datasets.

The first 10 seconds of recordings were discarded to ensure magnetization equilibration. Average white matter and cerebrospinal fluid signals were computed using restrictive masks from the DPARSFA toolbox (52), to avoid partial volume effects. The global signal was also computed, including voxels with a probabilistic grey matter signal larger than 0.025.

Individual voxelwise time courses of activity were subsequently detrended, and regressed for confounding variables including (a) linear and quadratic trends, (b) average white matter, cerebrospinal fluid and global signals, (c) a Discrete Cosine Transform basis with frequency cutoff 0.01 Hz, enabling high-pass temporal filtering, (d) the six head movement parameters obtained from realignment, and (e) one spike regressor per individual volume deemed corrupted by excessive framewise displacement (FD), computed according to (53). The FD threshold was set to 0.5 mm for all datasets.

After regression, the output functional volumes were spatially smoothed (5 mm full width at half maximum).

#### Determination of final sample sizes

##### Dataset 1

Although the full HCP dataset comprises more than 800 subjects, we focused our analyses on the *100 Unrelated* subjects only, for two reasons. First, in doing so, we could avoid the presence of any family relationships to correct for in the data. Second, a power analysis indicated that at α=0.05 and 80% power, a medium effect size of 0.3 (Cohen’s *d*) could be detected with a one-tailed *t*-test (to ascertain whether stability and differential identifiability would be significantly larger from 0) from *n*=72 samples. Furthermore, we did not conduct any more advanced analyses that would have necessitated a larger dataset (*e.g.*, multivariate brain/behavior associations). 54 female and 46 male participants (age: 29.1±3.7 years) were part of the analyzed dataset. Of these, 10 were excluded from the analyses due to excessive head movement (excessive framewise displacement [FD>0.5 mm; (53)] in more than 20% of frames in any of the four available resting-state scans), for a final sample size of *n*=90 subjects.

##### Dataset 2

The HCP-EP dataset comprises a total 123 early psychosis subjects with available neuroimaging and clinical data. A power analysis indicated that at α=0.05 and 80% power, a medium effect size of 0.3 (Pearson’s correlation coefficient) could be detected through correlational analysis (to relate symptoms of apathy and CB-VTA FC) from *n*=84 scans. Accordingly, we focused our investigations on the 106 subjects with early psychosis from the dataset for which all imaging and clinical data were available.

Amongst these, 7 were excluded due to excessive head movement (excessive framewise displacement [FD>0.5 mm; (53)] in more than 20% of frames in any of the four available resting-state scans), resulting in a final sample size of *n*=99 subjects for analyses (76 subjects with non-affective and 23 with affective psychosis).

Note that in additional stability and differential identifiability analyses (shown in **Supplementary Figure 3**), we also separately examined CB-VTA FC in *n*=30 healthy controls from the HCP-EP dataset satisfying similar quality criteria.

##### Dataset 3

27 patients and 7 controls at T_1_ and 12 patients and 9 controls at T_2_ were excluded during different stages of quality control of the imaging analysis. Reasons for exclusion were the lack of resting-state scans (8 subjects at T_1_ and 11 at T_2_), excessive movement (above threshold of 3 mm or more than 50% of frames with excessive FD: 18 subjects at T_1_ and 3 at T_2_), lacking the cerebellum in the field of view (8 subjects at T_1_ and 1 at T_2_), interrupting the study (5 subjects at T_2_), or epileptic seizure (1 at T_2_). Joint T_1_/T_2_ imaging data remained for *n*=19 HCs and *n*=23 SZ patients. In total, there were *n*=87 good quality scans from SZ patients (some contributing two, and others only one), and *n*=69 for HCs.

Note that this dataset was originally acquired for other purposes than the present investigations; as such, the available sample size was not determined through power analysis for this study, and all available data was leveraged.

#### Computation of CB-VTA FC

First, we separately quantified whole-brain normative FC with the right and with the left VTA, using the online *Neurosynth* platform (54; https://neurosynth.org/). The left and right VTA were obtained as their respective masks from the Automated Anatomical Labeling (AAL) atlas (55), and the coordinates input to *Neurosynth* were their centers of gravity. *Neurosynth* bases its computations on a set of 1,000 healthy individuals fully independent from the data analyzed therein. The two resulting maps were averaged together to yield a map representative of normative FC to the bilateral VTA (see **Supplementary Figure 1B**). We opted for performing analyses on the bilateral VTA rather than separately for each hemisphere for two reasons. First, the VTA is a small midline structure with limited inter-hemispheric separation at the spatial resolution of fMRI (2 mm isotropic); lateralized seeds risk capturing largely overlapping signal and reducing effective signal-to-noise ratio. Second, while structural connectivity work has identified lateralized cerebellum–midbrain pathways (11), our primary aim was to identify a generalized CB–VTA functional substrate suitable for TMS targeting, where coil focality and clinical translatability favor bilateral aggregation.

Second, we constructed the time course reflective of VTA activity as a weighted averaging across the activity time courses of all subcortical and cortical voxels (using FC of voxels to the bilateral VTA as weights; *seed map* approach), rather than simply averaging across only VTA voxels (*seed-only* approach). This enables to boost the signal-to-noise ratio of subject-level analyses, as the averaging is conducted over a much larger number of voxels, and has been recommended in recent studies (20,21). In our case, there were 192 VTA voxels, against a total of 139,942 (sub)cortical voxels. In dedicated supplementary investigations, we explicitly confirmed that in the context of our application, individual features of CB-VTA FC were captured more clearly with the *seed map* approach (see **Supplementary Methods – Constructing the time course of VTA activity** and **Supplementary Figure 2**). Note that the cerebellum target region was excluded from the voxels averaged over.

Given our focus on clinical application through TMS, we restricted the target region to TMS-accessible cerebellar sub-regions (*i.e.*, the posterior cerebellum), defined as region indices 8 to 19 from the Diedrichsen atlas (56; see **Supplementary Figure 1A**). This included the left/vermis/right Crus I, left/vermis/right Crus II, and left/vermis/right Lobules VIIB and VIIIA, for a total of 10,366 voxels.

Functional connectivity was computed as Pearson’s correlation coefficient, including only the time points that were not scrubbed out (FD<0.5 mm).

For Datasets 1 and 2, two separate pairs of scans were available. For analyses pertaining to stability and differential identifiability of CB-VTA FC, scans from each pair were concatenated to allow examination of these features across timescales spanning minutes (Dataset 2) to a few days (Dataset 1). The VTA activity time course was temporally z-scored for each scan beforehand, to further mitigate potential remaining cross-scan differences. Cerebellar activity was similarly temporally z-scored for each scan prior to concatenation. For Dataset 2 analyses regarding associations with apathy, all four scans were similarly merged to obtain one CB-VTA FC map per subject.

#### Stability and differential identifiability of CB-VTA maps

We quantified stability and differential identifiability similarly for all datasets. In each case, two CB-VTA FC maps were available per subject, computed with a dataset-specific inter-scan interval; we refer to them as session 1 and session 2. Each map was vectorized, and spatial similarity (Pearson’s correlation coefficient) was computed across subjects and sessions, yielding an *S* x *S* similarity matrix, with *S* the number of subjects. The diagonal of the matrix contained spatial similarity values across both sessions from the same subject, while the off-diagonal elements instead reflected similarity values across the sessions from separate subjects.

Stability was taken as the average of diagonal elements (*i*.*e*., mean similarity between FC maps from the same subject). Higher stability indicates more reliable FC patterns over time (values closer to 1 reflect stronger reproducibility), whereas lower values reflect reduced within-subject consistency.

Differential identifiability was quantified as the difference between the mean diagonal and off-diagonal elements (57). Because off-diagonal elements represent the similarity between FC maps from different subjects, a higher differential identifiability therefore reflects a greater separation between within-subject and between-subject similarity, indicating more individually distinctive FC patterns.

We favoured differential identifiability over the intra-class coefficient (ICC), where individual values would be obtained per voxel, the averaging of which would preclude the examination of spatial cerebellar variations. Furthermore, the ICC would have suffered from the unbalanced number of classes (the subjects) compared to measurements per class (the two CB-VTA FC maps).

#### Association between CB-VTA FC and apathy

Voxelwise associations were computed between CB-VTA FC and apathy severity, as measured by the CAINS (Dataset 2, early psychosis) or BNSS (Dataset 3, chronic schizophrenia), across all 10,366 cerebellar voxels. Candidate covariates were screened separately in Datasets 2 and 3 to determine whether apathy severity differed across categorical covariates or was associated with continuous covariates. Covariates showing no detectable relationship with apathy were not retained in the voxelwise models, to preserve model parsimony and avoid unnecessary loss of degrees of freedom; antipsychotic medication dose was retained in both datasets because of its direct clinical relevance. Statistical correction procedures are detailed in the **Statistical testing** subsection below.

In Dataset 2, psychosis subtype (affective versus non-affective) and antipsychotic dosage (quantified as mg of chlorpromazine equivalents) were retained as covariates in the model. Other candidate covariates were screened to determine whether they were associated with apathy severity. Apathy as quantified by the CAINS score did not differ across gender (Wilcoxon’s rank-sum test, female [0] versus male [1]: *z=*-0.67, *r*=0.07, *p*=0.5) and race (Wilcoxon’s rank-sum test, white [1] versus black [2]: *z*=-0.22, *r*=0.02, *p*=0.82; white [1] versus others [3]: *z*=0.02, *r*=0.003, *p*=0.98; black versus others: *z*=0.11, *r*=0.02, *p*=0.91), or as a function of age (Spearman’s correlation: *r*_97_=-0.07, *p*=0.49) and mean framewise displacement (Spearman’s correlation: *r*_97_=-0.03, *p*=0.76).

Consequently, voxelwise CB–VTA FC was modeled as a function of CAINS MAP apathy score as:

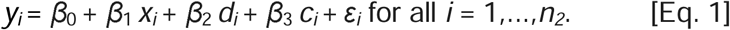

In the above formula, *y_i_* is CB-VTA FC at the location of interest for subject *i*, *x_i_* is the CAINS score, and *d_i_* and *c_i_* are the psychosis subtype and antipsychotic medication dosage for the same subject. ε*_i_*is the measurement error following a normal distribution of mean 0 and standard deviation σ, and *n*_2_=99 is the number of early psychosis patients analyzed in Dataset 2.

In Dataset 3, risperidone equivalent dosage (in mg) was available and was the only included covariate, given that apathy as quantified by the BNSS did not differ across gender (Wilcoxon’s rank-sum test, female [0] versus male [1]: *z=*-0.69, *r*=0.07, *p*=0.49) or as a function of age (Spearman’s correlation: *r*_85_=0.04, *p*=0.69) and mean framewise displacement (Spearman’s correlation: *r*_85_=0.16, *p*=0.17). Information on race was not recorded for this dataset.

The following mixed model was used to model voxelwise CB–VTA FC as a function of BNSS apathy score:

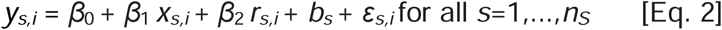

In the above, *y_s,i_*is CB-VTA FC at the location of interest for the *i*^th^ measurement of subject *s* (*i*=1 for subjects with one scan, and *i* ∈ [1,2] for subjects with two scans), *x_s,i_*and *r_s,i_* are the BNSS score and the risperidone equivalent medication dosage for the same subject and measurement, *b_s_*is the random effect for subject *s*, following a normal distribution of mean 0 and variance σ_b_ ^2^, ε is the measurement error following a normal distribution of mean 0 and standard deviation σ, and *n_S_*=65 is the number of subjects at hand (some with only 1 scan, and others with 2, for a total of 87 scans).

#### Meta-analysis of randomized, sham-controlled TMS trials

To evaluate whether the paravermal territories identified in Datasets 2 and 3 also contribute to therapeutic effects, we performed a secondary analysis of 39 sham-controlled TMS trials targeting negative symptoms in schizophrenia, including 867 patients receiving active stimulation and 757 receiving sham in a recently published meta-analysis (15), as an independent translational benchmark. Inclusion criteria were: (i) randomized, sham-controlled design; (ii) adult participants (≥18 years) with a schizophrenia spectrum disorder (schizophrenia, schizoaffective disorder, or psychosis); (iii) negative symptoms as a primary outcome; and (iv) stimulation parameters available. In this meta-analysis, clinical improvement on negative symptoms was reported for each trial as the standardized mean difference (SMD) in symptom change between the active and sham groups, corrected with Hedges’ g for small-sample bias. Full methodological details are provided in (15).

Using the published RCT dataset (15), we tested whether clinical improvement across trials was associated with the FC strength between each trial’s stimulation site and the cerebellum, and mapped where in the cerebellum this relationship emerged. We therefore focused on those trials with enough targeting information to localize the stimulation site in standardized MNI space. This yielded 39 trials with localizable stimulation sites. The remaining trials for which the stimulation site could not be reliably mapped were excluded from this secondary analysis. Across the retained trials, stimulation sites spanned the cerebellum as well as the left and right dorsolateral prefrontal cortex, right inferior parietal cortex, and left temporoparietal region, and included both high- and low- frequency TMS protocols. Study-level descriptions, derived MNI coordinates, stimulation parameters, and effect sizes are provided in **Supplementary Table 3**.

For each retained study, the stimulation sites were converted to MNI space; when provided as EEG landmarks, sites were mapped to MNI surface coordinates (58). Each stimulation site coordinate was then used as a seed in the *Neurosynth Locations* tool (https://neurosynth.org/locations/) a coordinate-based — rather than term-based — seed function that returns a whole-brain normative resting-state functional connectivity map computed from an independent reference sample of approximately 1,000 healthy adults. From each trial’s whole-brain map, we then extracted the cerebellar voxels (defined identically to the target region in our primary analyses, Diedrichsen atlas regions 8–19), yielding one normative cerebellar FC map per trial. Voxelwise associations between trial-level clinical improvement and normative cerebellar FC were then tested across the cerebellar mask, with stimulation region, stimulation frequency, and their interaction included as covariates; statistical inference is described below.

#### Statistical testing

##### Stability and differential identifiability

For each population of subjects at hand, stability was quantified as Pearson’s correlation coefficient of within-subject cross-scan similarity (*i.e.*, session 1 compared to session 2 of the *same* subject), averaged across subjects. To quantify differential identifiability, Pearson’s correlation coefficient of cross-subject cross-scan similarity (*i.e.*, session 1 compared to session 2 of a *different* subject) was computed and averaged across all subject pairs. This quantity was subtracted to stability to yield differential identifiability.

Significance for stability and differential identifiability was assessed by non-parametric permutation testing, randomly shifting subject labels for session 1 data 100,000 times and using the resulting null distributions. Robustness of the measures was examined with bootstrapping, selecting 80% of the data 100,000 times to construct a bootstrapped confidence interval.

##### Associations between CB-VTA FC and apathy

Associations between apathy scores and candidate covariates were examined with a Wilcoxon rank-sum test for discrete variables (computing the effect size as the standardized test statistic divided by the square root of the total number of paired observations), and Spearman’s correlation coefficient for continuous variables.

For Dataset 2, the generalized linear model described in **Eq. 1** was applied to cerebellar voxels with significance defined as *p*<0.05 (two-tailed), corrected for multiple comparisons using a Freedman-Lane non-parametric cluster-based family-wise error (FWE) procedure with 100,000 permutations, a cluster-forming threshold of p_c_=0.01, 26-connected clusters, implemented in Python’s *nilearn*. One cluster survived correction, in which more negative FC values were associated with lower CAINS scores (**Figure 4A**).

For Dataset 3, the mixed model described in **Eq. 2** was applied to cerebellar voxels with significance defined as *q*<0.05 (two-tailed) using false discovery rate (FDR) correction for multiple comparisons, given that permutation frameworks do not accommodate the mixed-effects structure. Two sizable clusters (164 and 158 voxels) survived correction, in which less positive FC values were associated with lower BNSS scores (**Figure 4B**). There were also four additional clusters of 1, 13, 2 and 4 voxels, but they were not analyzed further given their small size.

##### Meta-analysis

For each RCT, effect size was expressed as the standardized mean difference (SMD) with Hedge’s correction. Voxelwise associations between SMD and normative cerebellar FC were tested using the same two-tailed Freedman-Lane cluster-based FWE approach, with 100,000 permutations, a cluster-forming threshold of p_C_=0.05, 26-connected clusters in Python’s *nilearn*.

#### Software and data availability

Unless otherwise specified, all the analyses were performed using MATLAB (The MathWorks, Inc., California, United States of America; version 2024a).

Data from Datasets 1 and 2 can be accessed by accredited experimenters. Data from Dataset 3 are available upon reasonable request, subject to data-sharing agreements and within the limits of laboratory resources.

## Acknowledgement

We acknowledge the support and expertise of the MRI Platform at the Fondation Campus Biotech Geneva, co-founded and supported by the École Polytechnique Fédérale de Lausanne (EPFL), the University of Geneva (UNIGE), and the Geneva University Hospitals (HUG).

TAWB and LS contributed to formal analysis and visualization. TAWB and IB contributed to writing the original draft of the article. TAWB and IB contributed to methodology. TAWB, LS, HV, DVDV, SK, HC and IB contributed to the review and editing of the article. HV, SK, HC and IB contributed to data acquisition. IB contributed to conceptualization, hypothesis formulation and questions, supervision, resources, project administration, and funding acquisition.

## Funding

This work was supported by a Leenaards Foundation Grant (Translational Science Prize 2023 [to IB]), Projets recherche et développement (PRD) funds (Grant No. 22-2020-I [to IB]), a Swiss National Science Foundation Grant (No. 169783 [to SK]), and an excellence scholarship of the Swiss Government (ESKAS) awarded to LS.

## Conflict of interests

IB is the inventor, and TAWB and IB are named contributors on a pending patent related to methods of identifying targets for transcranial magnetic stimulation via CB-VTA functional connectivity. All other authors report no conflicts of interest.

## Notes

### Competing Interest Statement

Indrit Bègue is the inventor, and Thomas Bolton and Indrit Bègue are named contributors on a pending patent related to methods of identifying targets for transcranial magnetic stimulation via CB-VTA functional connectivity. All other authors report no conflicts of interest.

### Author Declarations

All participants provided informed consent under protocols approved by the Geneva Ethics Committee (CCER. BASEC IDs: 2017-01765, 2020-02169).

