## Supplementary Materials for "Cerebellum-ventral tegmental connectivity as a mechanism-informed target for apathy in schizophrenia"

### Supplementary Methods

#### Constructing the time course of VTA activity

In this study, we generate the time course reflective of VTA activity as a weighted averaging across all (sub)cortical voxels (using functional connectivity [FC] of voxels to the bilateral VTA as weights; *seed map* approach), rather than simply averaging across only VTA voxels (*seed-only* approach). This enables to boost the signal-to-noise ratio of subject-level analyses, as the averaging is conducted over a much larger amount of voxels.

Fox and colleagues (1) were the first to remark the importance of this step in the context of personalized TMS approaches. They were concerned with locating the maximal subgenual cingulate (SGC)-to-dorsolateral prefrontal cortex (DLPFC) anti-correlation in individual subjects, in the assessment of TMS benefits for major depressive disorder. Thus, the DLPFC was their target region, while the SGC was their seed region. In their hands, a *seed map* of subgenual FC obtained on a large independent cohort of 1000 subjects proved superior.

To provide more details, in Figure 4 from their study, they compared the similarity of DLPFC FC within individual subjects (imaged on two separate days, thus yielding *session 1* and *session 2* scans) versus across distinct individuals when (A) averaging over SGC voxels only (*seed-only* approach), or (B) conducting weighted averaging across all (sub)cortical voxels (except the target DLPFC region; *seed map* approach). Similarity values became much stronger in the latter case, evidencing a better ability to capture individual FC patterns. Furthermore, FC within subjects also became larger than FC between a subject and the “group FC map”, which was not the case in the seed-only case. This provided clear evidence that the subtleties of individual FC patterns can only be truly extracted with the seed map approach.

The authors also used session 1 scans, or the group FC map, to pinpoint the DLPFC focus of maximal anti-correlation with the SGC, at different spatial resolutions ranging from 30 mm to 1 mm (see their Figure 6). The anti-correlation was then quantified on the second, separate scan 2. Anti-correlations were much more marked using the seed map approach, and there was also a more tangible increase in anti-correlation strength from using a group-based to an individualized target in this case.

More recently, Cash *et al*. (2) also evidenced the superiority of the seed map approach compared to the simpler seed-only one. Using 14 or 28 min of recordings, they showed (see their Figure 4) that the distance between the exact spots of maximal SGC-DLPFC anti-correlation from a scan to the next was consistently lower in the seed map case, regardless of the specifics of the algorithm used to infer the optimal spot from the DLPFC FC map. Furthermore, the inter-/intra-individual distance ratios were also higher, demonstrating the superiority of the seed map approach in a personalization setting.

In light of these observations, we expected similar outcomes in the context of CB-VTA FC, as also supported by the large acknowledged inter-individual variability in cerebellar FC territories (3). In our case, there were 192 VTA voxels, against a total of 139,942 (sub)cortical voxels. Note that the cerebellum, our target region, was always excluded from the voxels averaged over.

**Supplementary Figure 2** shows the results of similarity assessments on Dataset 2 (HCP-EP, on a subset of *n*=46 subjects) when performed with the *seed map* approach (as in the main material) as compared to the *seed-only* approach. Evidently, the seed-only approach fails to adequately capture individual FC features, while the seed map approach can do so from using roughly 5 min of data. Stability and uniqueness are, expectedly, consistently larger in the seed map approach case, also stabilizing from roughly 5 min of data used onwards.

In short, our results show the superiority of the seed map approach over the seed-only one, and also demonstrate high stability and sufficient inter-individual variability for personalization as long as 5 min of data or more is used.

#### Complementary information on meta-analysis of randomized, sham-controlled TMS trials

This analysis builds on the published meta-analysis results in *“Mapping Symptom-General and Symptom-Specific Targets for Transcranial Magnetic Stimulation in Schizophrenia: An Electric-Field Modeling Meta-Analysis”* (4) by focusing specifically on randomized, sham-controlled trials of TMS for negative symptoms in schizophrenia.

Eligible studies were randomized, sham-controlled trials of TMS in adults with schizophrenia spectrum disorders in which negative symptoms were the primary outcome assessed with validated rating scales. Trials were excluded if another intervention was initiated concurrently, if outcome data were insufficient to compute effect sizes, or if the stimulation site could not be reliably localized in standardized space. The underlying systematic review was registered on PROSPERO (https://www.crd.york.ac.uk/prospero/display_record.php?RecordID=540178).

Of the 57 RCTs summarized in the forest plot of effect sizes for negative symptoms (Figure S3 in the Supplementary Materials of the original article), 39 studies (867 patients receiving active stimulation; 757 receiving sham) provided sufficient information on stimulation targets to be retained for voxel-wise connectivity analysis, while 18 were excluded because the TMS site could not be mapped to MNI or EEG coordinates. The excluded studies were: Bai *et al*. (2015), Bodén *et al*. (2021), Jin *et al*. (2023), Li *et al*. (2016), Liu *et al*. (2008), Ma *et al*. (2016), Prikryl *et al*. (2012), Wang *et al*. (2015, 2020), Xu *et al*. (2006, 2015), Zhang *et al*. (2015), Zhao *et al*. (2014a, 2014b, 2014c), and Zheng *et al*. (2012a, 2012b, 2012c).

For the retained studies, stimulation sites were transformed into MNI space, and these coordinates were then used to generate seed-based connectivity maps from *Neurosynth*. This enabled voxel-wise correlations between study-level standardized mean differences (effect sizes) and connectivity patterns, isolating cerebellar networks whose coupling to TMS targets predicted improvements in negative symptoms.

**Supplementary Table 3: Study overview and stimulation parameters for the secondary analysis on randomized, sham-controlled TMS trials targeting negative symptoms in schizophrenia.**

| **Study ID** | **Study** | **Total**  **sham** | **Total**  **treatment** | **Effect size SMD** | **Standard error** | **Stimulation site** | **MNI coordinate** | **Frequency** |
| --- | --- | --- | --- | --- | --- | --- | --- | --- |
| **1** | Bais et al 2014 | 16 | 31 | -0.007724826 | 0.30782901 | T3P3 | [-66, -52, 16] | Low |
| **2** | Barr et al 2012 | 12 | 13 | -0.008458215 | 0.400322172 | B_DLPFC | [-50, 30, 36] and [50, 30, 36] | High |
| **3** | Basavaraju et al 2021 | 30 | 30 | -0.921608364 | 0.271559723 | CB | [0, -78, -34] | High |
| **4** | Bation et al 2021 | 10 | 12 | 1.130828417 | 0.460864789 | L_DLPFC | [-35, 34, 45] | High |
| **5** | Brady et al 2019 | 3 | 8 | 2.148383449 | 0.817392884 | CB | [0, -78, -34] | High |
| **6** | Chauhan et al 2020 | 17 | 19 | -0.03311905 | 0.333871746 | CB | [0, -78, -34] | High |
| **7** | Chen et al 2011 | 19 | 23 | 0.48636665 | 0.314524945 | L_DLPFC | [-36, 49, 32] | High |
| **8** | Chibarro et al 2005 | 8 | 8 | 1.107915731 | 0.536990377 | T3P3 | [-66, -52, 16] | Low |
| **9** | Cordes et al 2010 | 13 | 12 | 0.592470616 | 0.408994913 | L_DLPFC | [-38, 25, 49] | High |
| **10** | De Jesus et al 2011 | 9 | 8 | 0.331120111 | 0.489219616 | T3P3 | [-66, -52, 16] | Low |
| **11** | Dlabac-de Lange et al 2014 | 16 | 16 | 0.335667096 | 0.356034417 | B_DLPFC | [-36, 49, 32] and [40, 48, 32] | High |
| **12** | Dollfus et al 2018 | 33 | 26 | 0.056122365 | 0.262280882 | T3P3 | [-66, -52, 16] | High |
| **13** | Duan et al 2013 | 20 | 21 | 0.751776665 | 0.32328214 | L_DLPFC | [-36, 49, 32] | High |
| **14** | Fitzgerald et al 2008 | 8 | 12 | 0.11208191 | 0.456779369 | B_DLPFC | [-38, 25, 49] and [38, 25, 49] | High |
| **15** | Garg et al 2016 | 20 | 20 | 0.169582578 | 0.316795641 | CB | [0, -78, -34] | High |
| **16** | Hajak et al 2004 | 10 | 10 | 0.991659336 | 0.473903688 | L_DLPFC | [-38, 25, 49] | High |
| **17** | Holi et al 2004 | 11 | 11 | -0.453859298 | 0.431856149 | L_DLPFC | [-38, 25, 49] | High |
| **18** | Huang et al 2016 | 18 | 19 | 0.028810445 | 0.328935178 | L_DLPFC | [-38, 25, 49] | High |
| **19** | Klein et al 1999 | 15 | 16 | -0.025812292 | 0.359412594 | R_DLPFC | [-35, 34, 45] | Low |
| **20** | Kos et al 2024 | 16 | 32 | -0.043280439 | 0.30621808 | R_DLPFC | [40, 48, 32] | High |
| **21** | Kumar 2020 | 50 | 50 | 0.435236512 | 0.202354032 | L_DLPFC | [-36, 30, 47] | High |
| **22** | Mogg et al 2007 | 9 | 8 | 0.269224767 | 0.488101356 | L_DLPFC | [-38, 25, 49] | High |
| **23** | Paillere et al 2016 | 12 | 15 | -0.09091621 | 0.387495896 | T3P3 | [-66, -52, 16] | Low |
| **24** | Pan et al 2021 | 20 | 16 | -0.034810885 | 0.335435285 | L_DLPFC | [-36, 49, 32] | High |
| **25** | Prikryl et al 2013 | 17 | 23 | 1.290703861 | 0.350892784 | L_DLPFC | [-41, 16, 54] | High |
| **26** | Prikryl et al 2014 | 17 | 18 | 0.302016394 | 0.340120774 | L_DLPFC | [-38, 25, 49] | High |
| **27** | Quan et al 2015 | 39 | 78 | 0.356169943 | 0.197493449 | L_DLPFC | [-38, 25, 49] | High |
| **28** | Rabany et al 2014 | 10 | 20 | 1.326044617 | 0.423446068 | L_DLPFC | [-36, 30, 47] | High |
| **29** | Ren et al 2011 | 11 | 12 | 0.60269731 | 0.426777499 | B_DLPFC | [-36, 49, 32] and [40, 48, 32] | High |
| **30** | Saba et al 2006 | 8 | 8 | -0.651117807 | 0.513077553 | L_DLPFC | [-50, -42, 19] | Low |
| **31** | Singh et al 2020 | 15 | 15 | 0.146295539 | 0.365636486 | L_DLPFC | [-38, 25, 49] | High |
| **32** | Tikka et al 2017 | 7 | 8 | 0.188618325 | 0.518693588 | R_INFERIOR_PARIETAL | [30, -56, 20] | High |
| **33** | Wen et al 2021 | 26 | 26 | -0.306831345 | 0.278977278 | L_DLPFC | [-32, 26, 52] | High |
| **34** | Wobrock et al 2015 | 81 | 76 | 0.094145588 | 0.159786736 | L_DLPFC | [-36, 49, 32] | High |
| **35** | Xiu et al 2020a | 30 | 32 | -0.198769586 | 0.254758625 | L_DLPFC | [-40, 40, 36] | High |
| **36** | Xiu et al 2020b | 30 | 35 | 0.05031629 | 0.248845809 | L_DLPFC | [-40, 40, 36] | High |
| **37** | Zhang et al 2010 | 12 | 15 | 0.635408084 | 0.396833378 | L_DLPFC | [-38, 25, 49] | High |
| **38** | Zhu et al 2021 | 32 | 32 | 0.21346691 | 0.250710991 | CB | [0, -78, -34] | High |
| **39** | Zhuo et al 2019 | 27 | 33 | 0.421436484 | 0.26233593 | L_DLPFC | [-36, 49, 32] | High |
| Abbreviations: T3P3 temporoparietal, B_DLPFC bilateral dorsolateral prefrontal cortex, CB cerebellum, L_DLPFC left dorsolateral prefrontal cortex, R_DLPFC right dorsolateral prefrontal cortex, R_INFERIOR_PARIETAL right inferior parietal | | | | | | | | |

### Supplementary Results

### Association of stability and differential identifiability with demographic and clinical measures

On all datasets, we assessed whether stability or differential identifiability (DI) were related to age, gender, race (when available, for Datasets 1 and 2), or psychosis subtype (for Dataset 2). Links with discrete/continuous variables were quantified with Wilcoxon’s rank-sum test and Spearman’s correlation, respectively.

For stability, our data vector was the diagonal of the CB-VTA FC cross-session similarity matrix (*i.e.*, all individual subject-specific cross-session similarity values). For DI, for each subject, we computed the average similarity of session *i* or *j* to all other subjects’ sessions *j* or *i.* This yielded two data vectors (one per cross-session comparison), which we averaged into our measure of interest. As such, one can view the output metric as reflective of how similar a subject’s CB-VTA FC map is to that of others on average.

For Dataset 1 (*n*=90 healthy controls), there was no association between stability and age (*r*_88_=0.1, *p*=0.37) or race (white [1] versus black [2]: *z*=-0.84, *r*=0.09, *p*=0.4; white [1] versus others [3]: *z*=1.06, *r*=0.12, *p*=0.29; black [2] versus others [3]: *z*=1.1, *r*=0.23, *p*=0.27). There was a mildly significant association of stability with gender (female [0] versus male [1]: *z*=2.09, *r*=0.22, *p*=0.037). There was no association between DI and age (*r*_88_=-0.02, *p*=0.87) or race (white [1] versus black [2]: *z*=-0.18, *r*=0.02, *p*=0.86; white [1] versus others [3]: *z*=0.58, *r*=0.07, *p*=0.56; black [2] versus others [3]: *z*=0.47, *r*=0.1, *p*=0.64), but there was an association of DI with gender (female [0] versus male [1]: *z*=3.44, *r*=0.36, *p*=5.75·10^-4^), albeit uncorrected for multiple comparisons.

For Dataset 2 (*n*=99 early psychosis subjects), there was no association between stability and age (*r*_97_=0.004, *p*=0.97), gender (female [0] versus male [1]: *z*=0.29, *r*=0.03, *p*=0.78), race (white [1] versus black [2]: *z*=1.91, *r*=0.2, *p*=0.06; white [1] versus others [3]: *z*=-0.6, *r*=0.08, *p*=0.55; black [2] versus others [3]: *z*=-1.02, *r*=0.16, *p*=0.31) or psychosis subtype (non-affective [1] versus affective [2]: *z*=-0.51, *r*=0.05, *p*=0.61). There was no association between DI and age (*r*_88_=-0.09, *p*=0.38), gender (female [0] versus male [1]: *z*=-0.54, *r*=0.05, *p*=0.59), or psychosis subtype (non-affective [1] versus affective [2]: *z*=0.53, *r*=0.05, *p*=0.59). Regarding race, while DI did not significantly differ between white and others (*z*=-1.13, *r*=0.16, *p*=0.26) or black and others (*z*=-1.55, *r*=0.25, *p*=0.12), white subjects tended to be more similar to the average than black subjects (*z*=2.66, *r*=0.27, *p*=0.008).

For Dataset 3, in healthy controls (*n*=19), neither stability nor DI were associated with gender (female [0] versus male [1], respectively: *z*=1.25, *r*=0.29, *p*=0.21 and *z*=0.27, *r*=0.06, *p*=0.79). However, there were significant positive associations with age (respectively: *r*_17_=0.5, *p*=0.03 and *r*_17_=0.68, *p*=0.0015), so that older subjects showed more stable CB-VTA FC maps, and also tended to be more similar to the average (*i.e.*, less distinctive). In chronic schizophrenia patients (*n*=23), there was no association of stability or DI with age (respectively: *r*_21_=-0.17, *p*=0.44 and *r*_21_=-0.22, *p*=0.32), and only a mildly significant one with gender (respectively: *z*=1.99, *r*=0.41, *p*=0.047 and *z*=1.99, *r*=0.41, *p*=0.047).

In short, no associations were consistent across all datasets, and taking the multiplicity of statistical testing into account, further more specific explorations should be conducted before any conclusion can be made.

### Supplementary Discussion

### Location of unraveled clusters

One can gain insight about the cerebellar clusters brought forward by our analyses by observing how their locations fare with respect to past research that subdivided the cerebellum into distinct subunits (**Supplementary Figure 7**). Considering the classical subdivision of the cerebellum in terms of resting-state networks (5, **Supplementary Figure 7A**), C_1_ lies within the default mode network (DMN), while C_A_ and C_B_ are primarily located within the antagonistic dorsal attention network (DAN). C_II_, meanwhile, is located at the intersection between the DMN, DAN and frontoparietal network (FPN). The high spatial variability across individuals in terms of exact network territories (3) precludes more detailed observations at this stage, but in future research leveraging sufficiently dense sampling, it will be interesting to determine how exactly subject-specific cerebellar network territories associate with apathy-related areas.

When scrutinized in terms of the second cerebellar functional gradient (which contrasts *task-unfocused* and *task-focused* cerebellar locations; 6, **Supplementary Figure 7B**), while C_1_ and C_I_ are located in task-unfocused cerebellar areas (blue underlay), C_A_ and C_B_ rather lie within task-focused ones (red underlay). This shows that apathy, and the impairments in goal-directed behavior and motivation that it yields, tightly relates to the balance between introspective and extrospective brain processing.

In a functional atlas generated from 7 large-scale task-based datasets (7, **Supplementary Figure 7C**), C_1_, C_I_ and C_II_ fall within “social-linguistic-spatial” areas, while C_A_ and C_B_ are rather found in “demand” areas. Collectively, these observations suggest that anti-correlation of the VTA to the cerebellum (as seen in C_1_) highlights an introspection-related, *task-unfocused* spot, while positive coupling between the VTA and the cerebellum would rather denote a location devoted to high-level executive functions (“demand”).

### Supplementary Figures

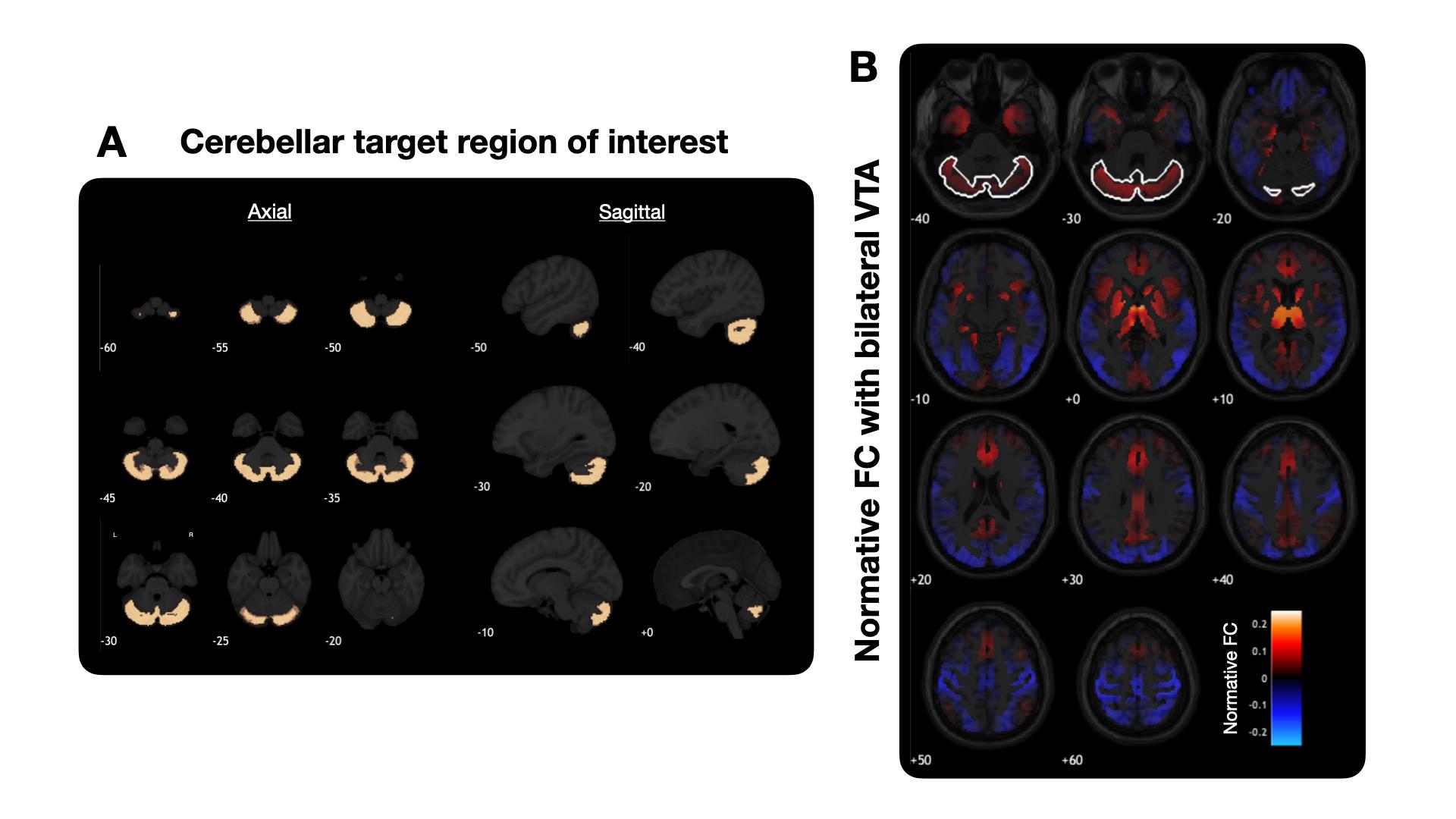

**Supplementary Figure 1: Target cerebellar region and normative functional connectivity to the bilateral ventral tegmental area.** **(A)** The cerebellar target region is salmon-coloured across selected axial and sagittal slices. A total of 10366 voxels are included. **(B)** Normative functional connectivity to the bilateral ventral tegmental area is shown across all (sub)cortical brain voxels. The white contour labels the cerebellar target region (whose values are *not* used for the computation of VTA activity).

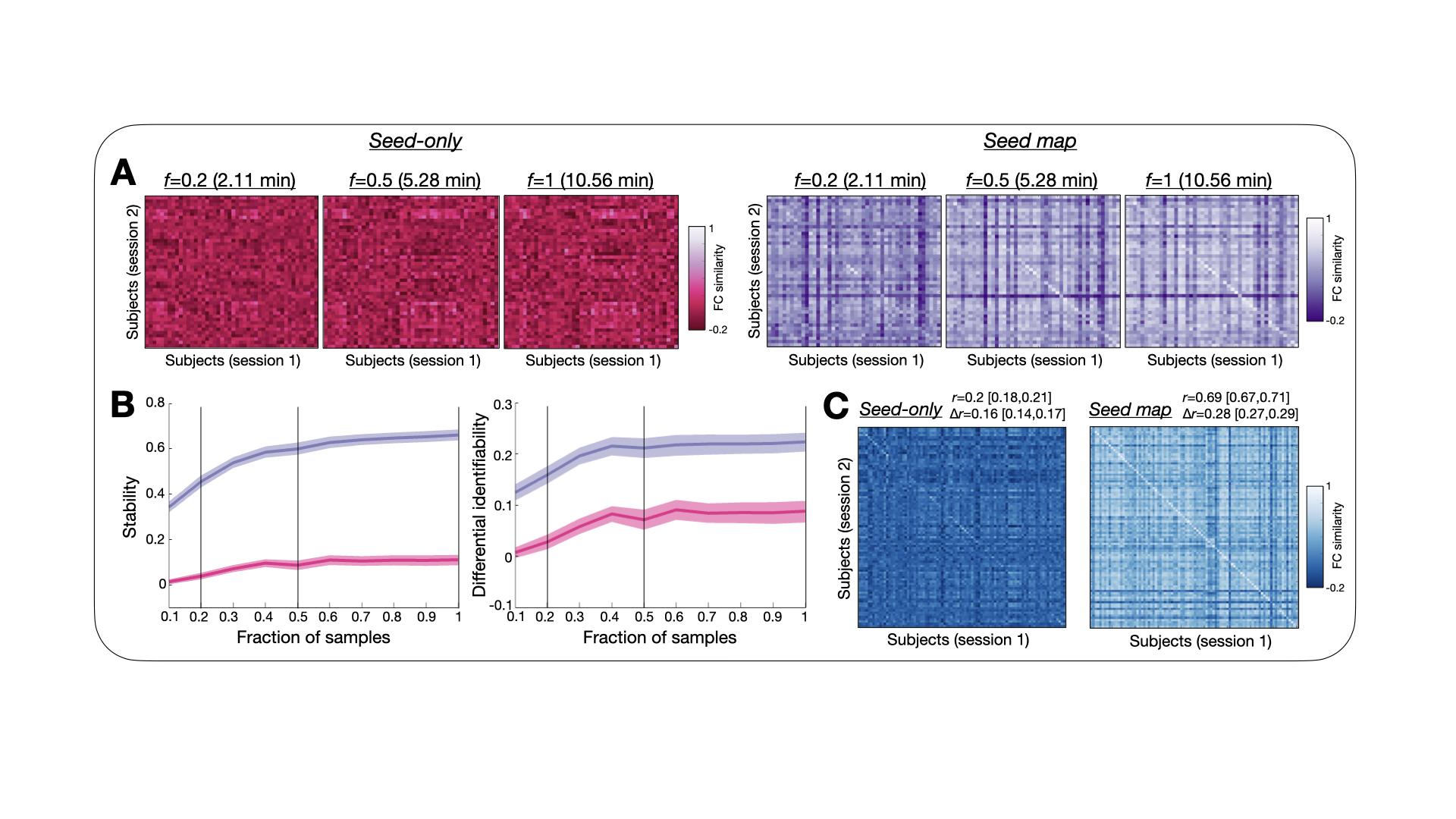

**Supplementary Figure 2: A seed map approach to compute VTA activity outperforms a seed-only counterpart. (A)** On a subset of *n*=46 subjects from Dataset 2 (HCP-EP), indicative cross-session similarity matrices when considering gradually longer recordings, using the *seed-only* (left) or *seed map* (right) approach to quantify VTA activity. The greater similarity between sessions from the same subject can only be seen with the seed map approach (higher diagonal than off-diagonal similarity values), from using 5 min of data onwards. **(B)** Accordingly, stability (left) and differential identifiability (right) are consistently larger in the seed map case, and stabilize from 5 min of data used onwards. **(C)** The better performance of the seed map approach is confirmed on Dataset 1 (HCP-YA, *n*=90), both in terms of stability and uniqueness (written down with their respective 95% confidence intervals).

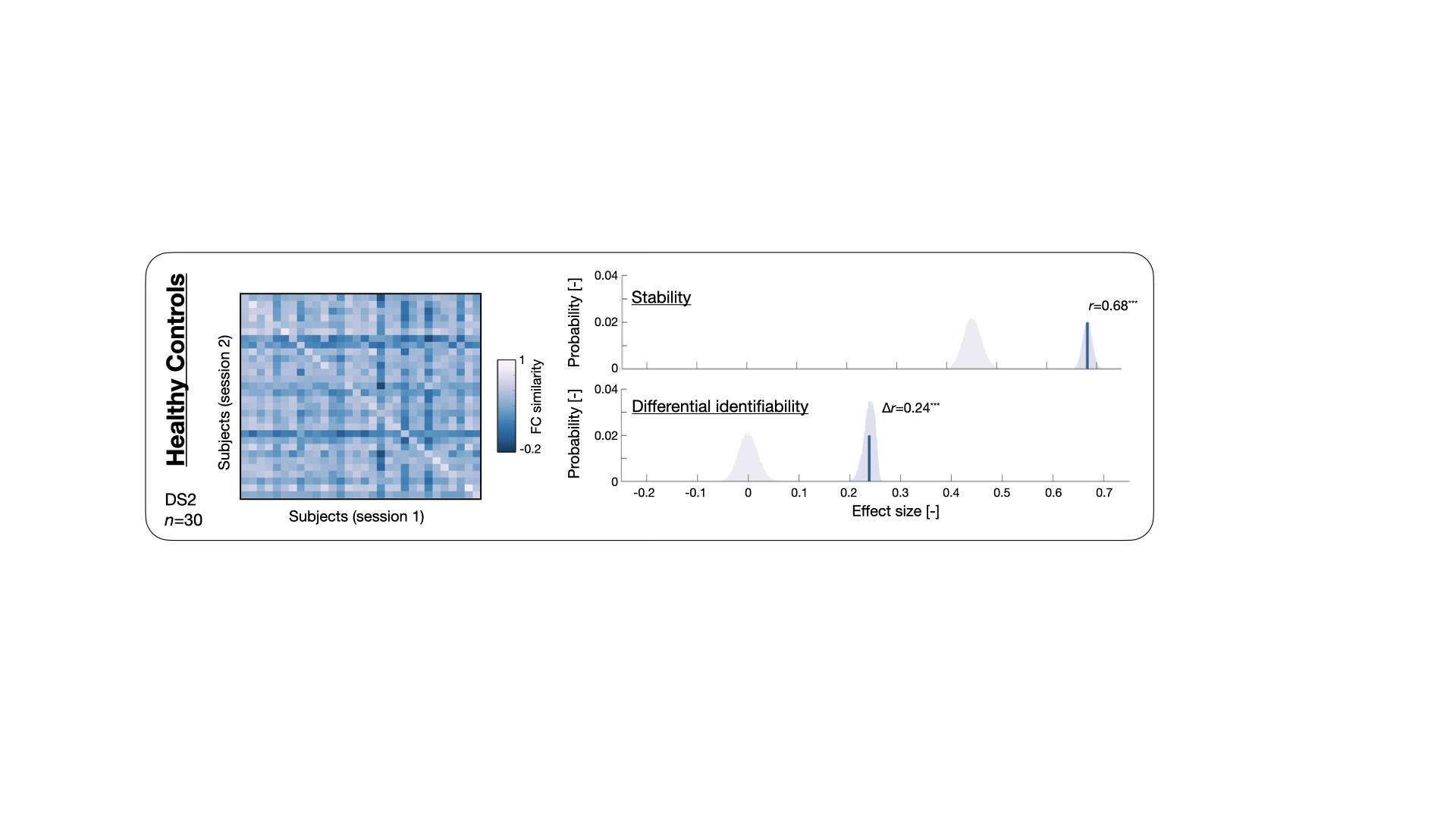

**Supplementary Figure 3: Cerebellum-ventral tegmental functional connectivity maps are stable and subject-specific in healthy subjects (HCP-EP dataset).** (*Left*) Similarity between FC maps at the population level (*n*=30). Diagonal/off-diagonal elements reflect cross-session similarity values within or across subjects, respectively. (*Right*) Stability (mean of diagonal similarity elements) was significant (top panel, *r*=0.68, *p*<10^-5^, light/dark blue null distribution/bootstrapped confidence interval), and so was differential identifiability (difference between mean [off-]diagonal similarity values; Δ*r=*0.24, *p*<10^-5^).

*Abbreviations:* FC: functional connectivity.

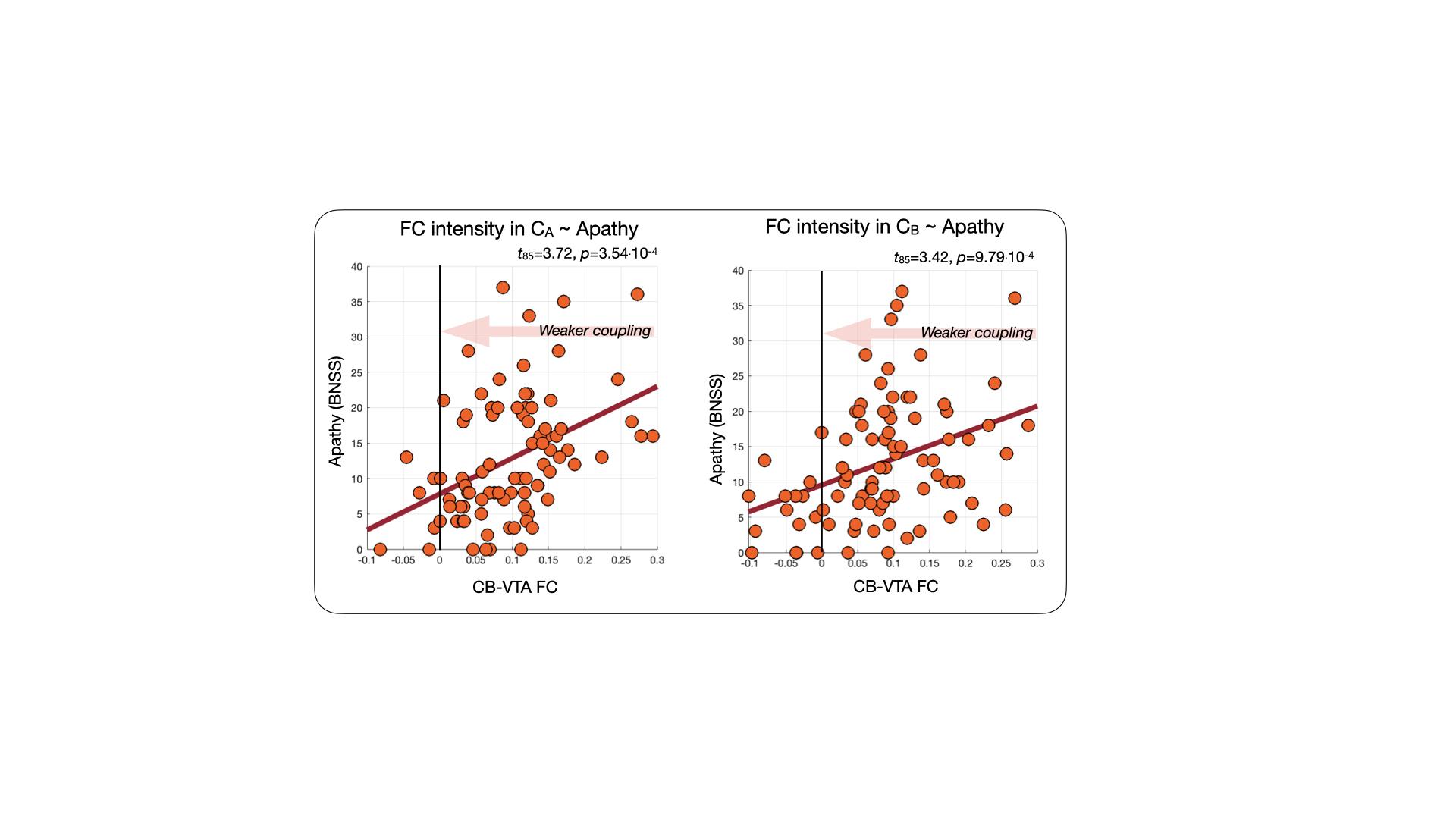

**Supplementary Figure 4: Cerebellum-ventral tegmental functional connectivity correlates with apathy in chronic schizophrenia patients (separate C_A_ and C_B_ analyses).** (*Left*) Association between C_A_ (top) or C_B_ (bottom) mean CB-VTA FC and apathy (respectively *t*_85_=3.72, *p*=3.54·10^-4^ and *t*_85_=3.42, *p*=9.79·10^-4^). (*Middle*) For C_A_ (top two plots) and C_B_ (bottom two plots), probability density function (top) and cumulative density function (CDF, bottom) for CB-VTA FC, color-coded for the severity of apathy. The middle plot shows *t*-statistics as a function of FC threshold for the association between CDF values and apathy within C_A_ (in dark red), C_B_ (in light red) or the whole cerebellar target region (in black). (*Right*) Relationship between CDF values and apathy scores at optimal FC thresholds (FC=0.06, *t*_85_=-4.04, *p*=1.16·10^-4^ for C_A_; FC=0.01, *t*_85_=-4.17, *p*=7.43·10^-5^ for C_B_). C_A_, C_B_: Clusters A and B, BNSS: Brief Negative Symptom Scale, CB-VTA FC: cerebellum-ventral tegmental area functional connectivity, N_voxels_: number of voxels, Cum. frac.: cumulative fraction.

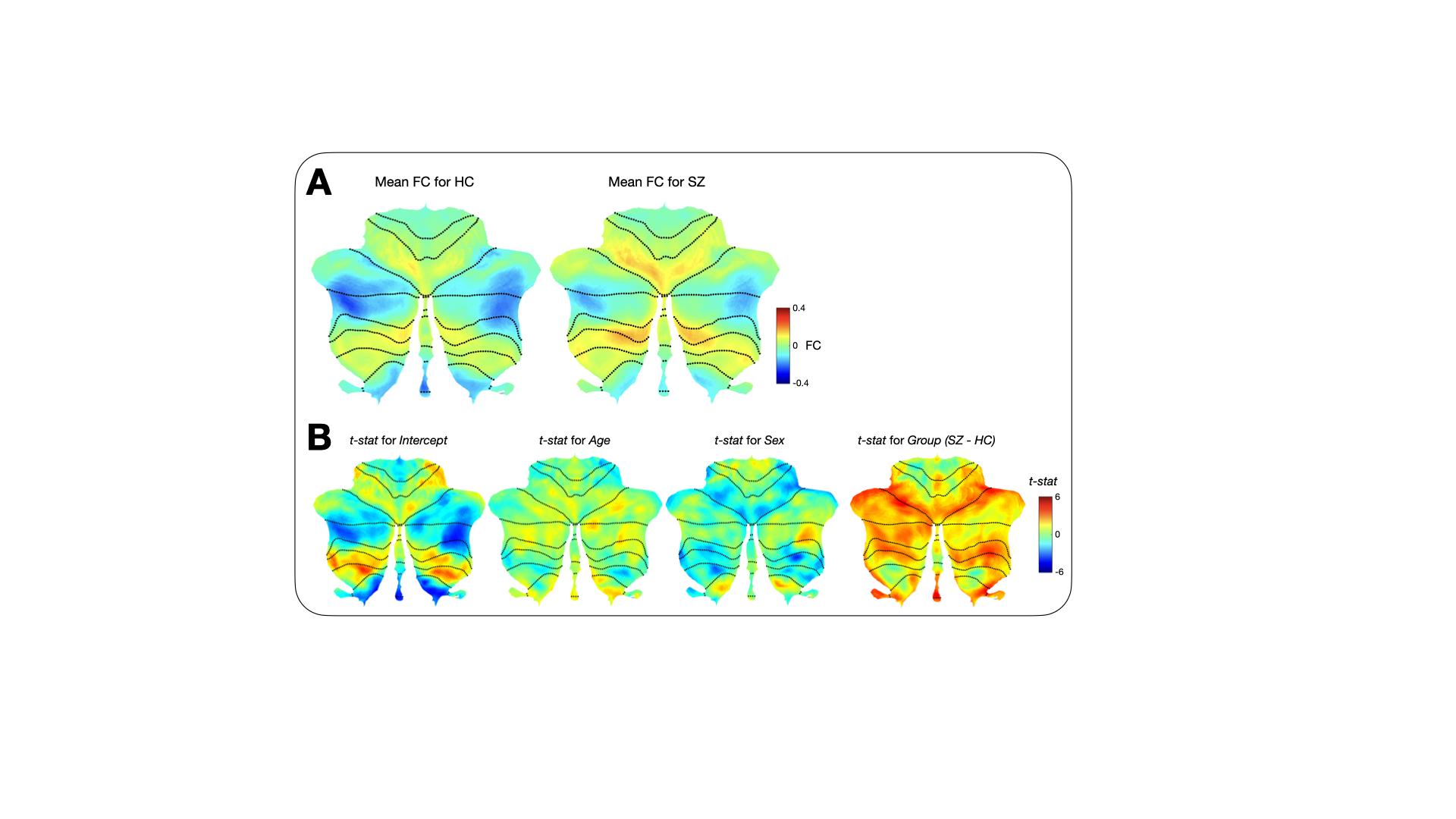

**Supplementary Figure 5: Group differences in cerebellum-VTA connectivity between healthy controls and chronic schizophrenia patients.** **(A)** For healthy controls (left, *n*=69 scans) and chronic schizophrenia patients (right, *n*=87 scans) from Dataset 3, average group-wise CB-VTA FC maps. **(B)** *t*-statistic maps for the intercept (left), age (middle left), sex (middle right) and group (right) effects upon mixed modelling of the patients group CB-VTA FC data according to FC ~ 1+Age+Sex+Group+(1|Subject ID). Note that the intercept map can be interpreted as the CB-VTA FC pattern seen in healthy controls after regressing out age and sex.

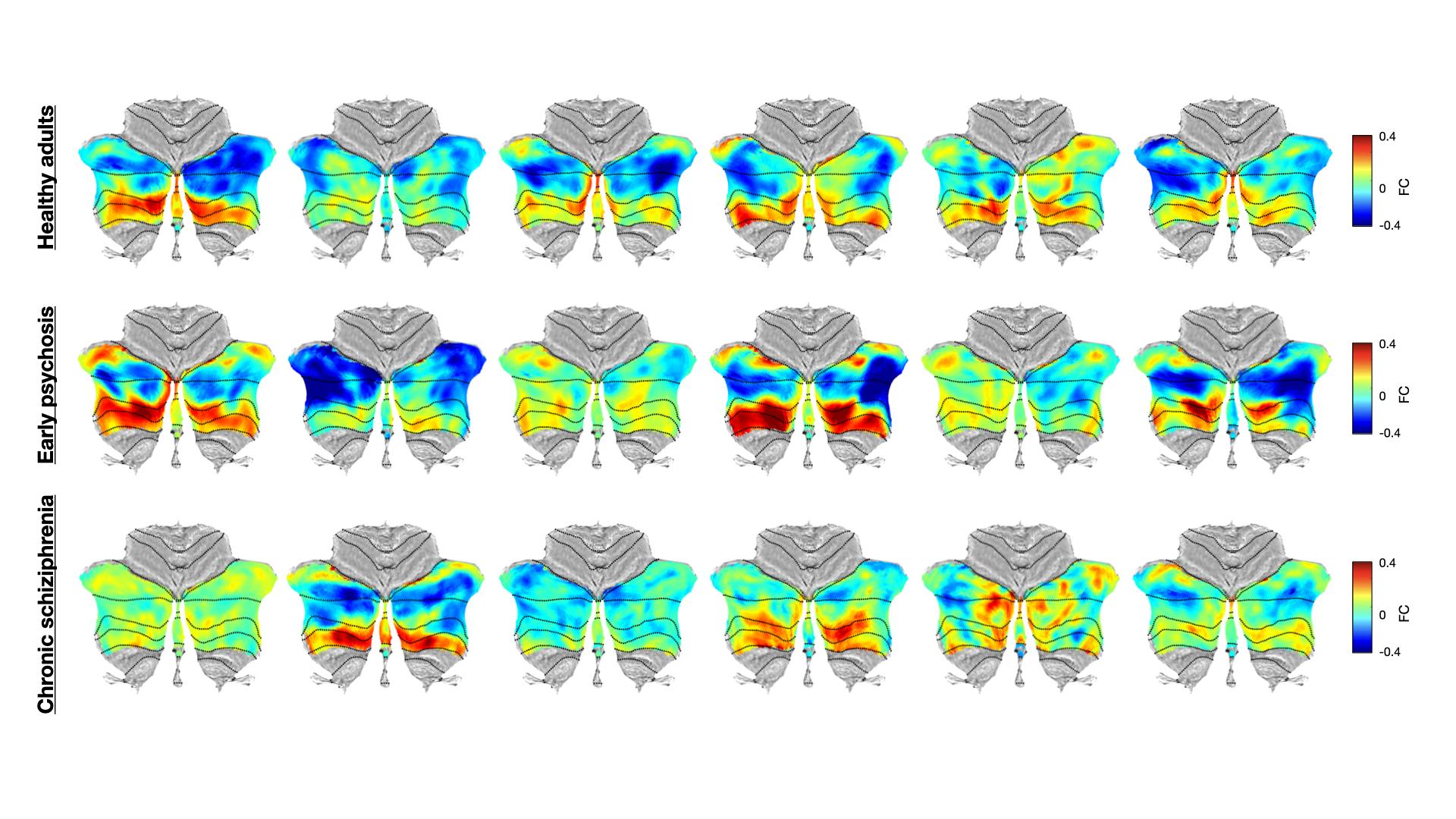

**Supplementary Figure 6: Characteristic CB-VTA FC maps across cohorts.** For healthy controls for Dataset 2 (top row), early psychosis subjects from the same dataset (middle row) and chronic schizophrenia patients from Dataset 3 (bottom row), six characteristic CB-VTA FC maps are shown to appreciate the extent and granularity of inter-subject variability. In each dataset case, clustering was performed on all available maps (correlation distance measure, *K*=6 clusters), and the most similar maps to each centroid were selected for display, to ensure that they are representative of the population.

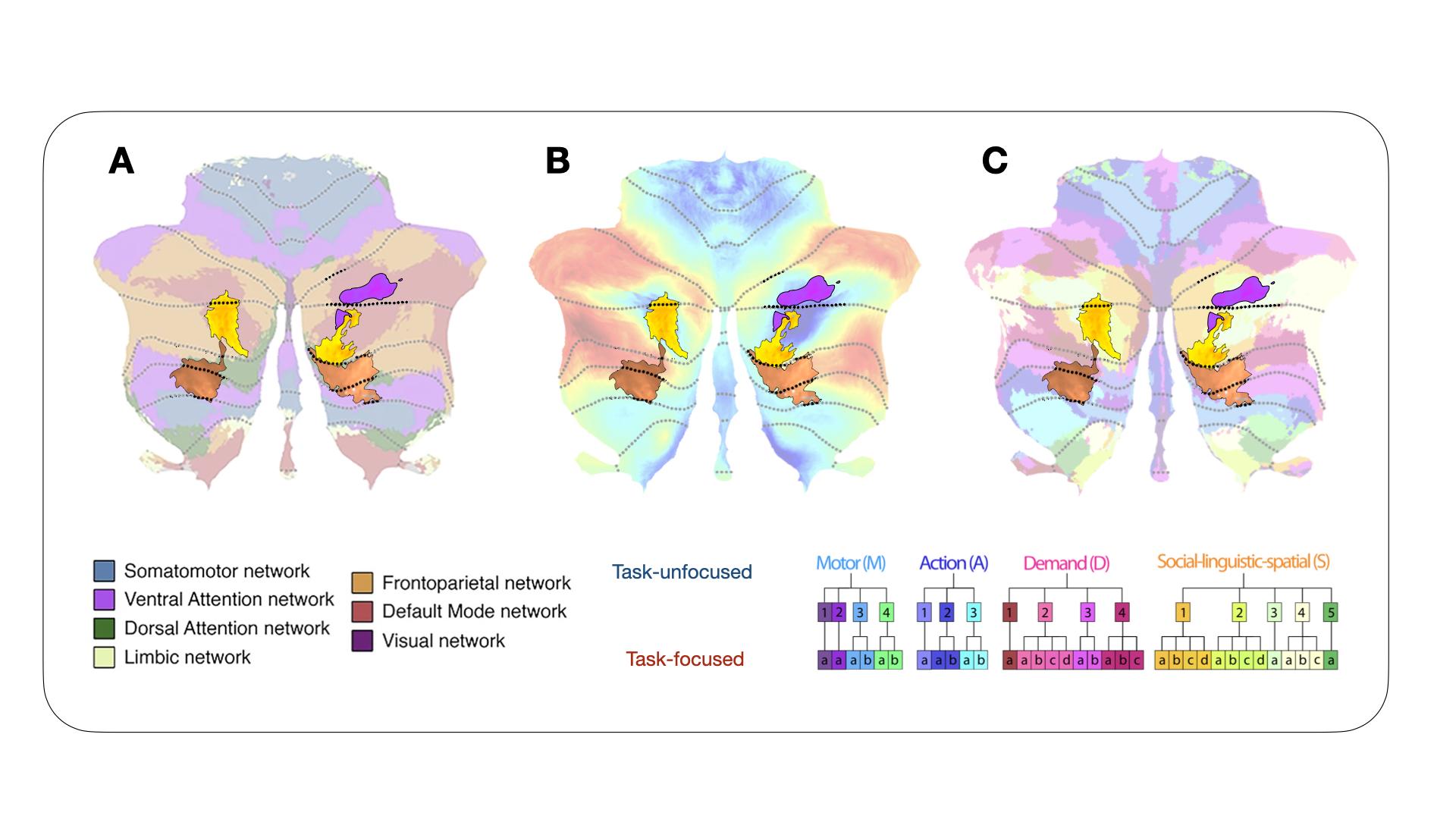

**Supplementary Figure 7: Location of apathy-related cerebellar clusters compared to past research.** C_1_ (purple), C_A_/C_B_ (copper), and C_I_/C_II_ (yellow) can be studied with respect to past cerebellar partitioning methodologies: into canonical resting-state networks **(A)** as per (5), into a task-unfocused to task-focused functional gradient **(B)** as per (6), or into a subdivision from a collection of task-based datasets **(C)** as per (7).
