## Supplementary Table 1 for "Cerebellum-ventral tegmental connectivity as a mechanism-informed target for apathy in schizophrenia"

**Supplementary Table 1. Inclusion and exclusion criteria for Dataset 1. Criteria are taken from Van Essen et al. (2013).**

| | |
|---|---|
| **Inclusion criteria** | |
| 1 | Age 22 to 35 at the time of telephone diagnostic interview (SSAGA) |
| 2 | Ability to give valid informed consent |
| **Exclusion criteria** | |
| 1 | Significant history of psychiatric disorder, substance abuse, neurological, or cardiovascular disease: (A) Participant report of diagnosis by a treating physician; or (B) Hospitalization for the condition for two days or longer; or (C) Pharmacologic or behavioral treatment by a cardiologist, psychiatrist, neurologist, or endocrinologist for a period of 12 months or longer, other than treatment for childhood-only ADHD |
| 2 | Two or more seizures after age 5 or a diagnosis of epilepsy |
| 3 | Any genetic disorder, such as cystic fibrosis or sickle cell disease |
| 4 | Multiple sclerosis, cerebral palsy, brain tumor or stroke |
| 5 | Any of the following head injuries: (A) Loss of consciousness for >30 minutes; or (B) Amnesia for >24 hours; or (C) Change in mental status for >24 hours; or (D) CT findings consistent with traumatic brain injury; or (E) Three or more concussive (mild) incidences of head injury |
| 6 | Premature birth (for twins, before 34 weeks; for non-twin siblings, before 37 weeks. If weeks unknown, less than 5 lbs. at birth for non-twins |
| 7 | Currently on chemotherapy or immunomodulatory agents, or history of radiation or chemotherapy that could affect the brain |
| 8 | Thyroid hormone treatment in the past month |
| 9 | Treatment for diabetes in the past month (other than gestational or diet-controlled diabetes) |
| 10 | Use of daily prescription medications for migraines in the past month |
| 11 | A score of 25 below on the Holstein Mini Mental State Exam (Holstein et al., 1975) on visit Day 1 |
| 12 | Moderate or severe claustrophobia |
| 13 | Pregnancy |
| 14 | Unsafe metal in the body |

| |
|---|
| **Inclusion criteria** |
| 1 |
| 2 |
| **Exclusion criteria** |
| 1 |
| 2 |
| 3 |
| 4 |
| 5 |
| 6 |
| 7 |
| 8 |
| 9 |
| 10 |
| 11 |
| 12 |
| 13 |
| 14 |
