## Supplementary Table 2 for "Cerebellum-ventral tegmental connectivity as a mechanism-informed target for apathy in schizophrenia"

**Supplementary Table 2. Inclusion and exclusion criteria for Dataset 2. Criteria are taken from Jacobs et al. (2025).**

| | |
|---|---|
| **Inclusion criteria** | |
| 1 | (A) DSM_V diagnosis of schizophrenia, schizophreniform disorder, schizoaffective disorder, psychosis not otherwise specified, delusional disorder, or brief psychotic disorder with onset within the past five years prior to study entry (non-affective psychosis); or (B) DSM-V diagnosis of major depression with psychosis (single and recurrent episodes) or bipolar disorder with psychosis (including most recent episode depressed and manic types) with onset within five years prior to study entry (affective psychosis) |
| 2 | 16 to 35 years of age at study entry |
| 3 | Male or female |
| 4 | Ability to provide informed consent or have a legal authorized representative or guardian |
| 5 | Outpatient |
| 6 | Fluent in English |
| 7 | Female subjects of childbearing age report that they are not pregnant and must test negative on a urine pregnancy test at the MRI visit |
| 8 | Willing to share de-identified data with the Connectome database |
| **Exclusion criteria** | |
| 1 | Substance-induced psychosis or psychotic disorder due to a medical condition |
| 2 | Known IQ less than 70 based on medical history |
| 3 | Subjects with known medical history of Human Immunodeficiency Virus positive (HIV+) status |
| 4 | Subjects with an active medical condition that affects brain or cognitive functioning (e.g., seizure disorder, epilepsy, head trauma, stroke, traumatic brain injury, significant loss of consciousness, or other neurological disorder) in the site principal investigator's opinion |
| 5 | Subjects with implanted pacemaker, medication pump, vagal stimulator, deep brain stimulator, TENS unit, ventriculoperitoneal shunt, or other contraindication to undergoing an MRI scan |
| 6 | Current severe substance use disorder in past 90 days (excluding caffeine and nicotine) |
| 7 | Electroconvulsive therapy treatment in past 12 months |
| 8 | Subjects considered a high risk for suicidal acts - active suicidal ideation as determined by clinical interview or any suicide attempt in 30 days prior to screening |
| 9 | Subjects who demonstrate overtly aggressive behavior or who are deemed to pose a substantial risk of danger in the investigator's opinion |

| |
|---|
| **Inclusion criteria** |
| 1 |
| 2 |
| 3 |
| 4 |
| 5 |
| 6 |
| 7 |
| 8 |
| **Exclusion criteria** |
| 1 |
| 2 |
| 3 |
| 4 |
| 5 |
| 6 |
| 7 |
| 8 |
| 9 |
